# Novel Genetic Loci Associated with Osteoarthritis in Multi-Ancestry Analyses in 484,374 Participants from MVP and the UK Biobank

**DOI:** 10.1101/2022.06.14.22276308

**Authors:** Merry-Lynn N. McDonald, Preeti Lakshman Kumar, Vinodh Srinivasasainagendra, Ashwathy Nair, Alison Rocco, Ava C. Wilson, Joe Chiles, Joshua Richman, Sarah A. Pinson, Richard Dennis, Vivek Jagadale, Cynthia Brown, Saiju Pyarajan, Hemant K. Tiwari, Marcas M. Bamman, Jasvinder A. Singh, the VA Million Veteran Program

**Affiliations:** Birmingham Veterans Affairs Healthcare System (BVAHS), Birmingham, AL, USA; Division of Pulmonary, Allergy and Critical Care Medicine, Department of Medicine, School of Medicine, University of Alabama at Birmingham (UAB), Birmingham, AL, USA; Department of Epidemiology, School of Public Health, UAB, Birmingham, AL, USA; Department of Genetics, School of Medicine, UAB, Birmingham, AL, USA; Department of Biostatistics, School of Public Health, UAB, Birmingham, AL, USA; Department of Surgery, School of Medicine, UAB, Birmingham, AL, USA; Central Arkansas Veterans Healthcare System (CAVHS), Little Rock, AR, USA; Department of Medicine, Louisiana State University Health Sciences Center, New Orleans, LA, USA; Veterans Affairs Boston Health Care (VABHC), Boston, MA, USA; Department of Cell, Developmental, and Integrative Biology, School of Medicine, UAB, Birmingham, AL, USA; Florida Institute for Human & Machine Cognition, Pensacola, FL, USA; Division of Rheumatology and Clinical Immunology, Department of Medicine at the School of Medicine, UAB, Birmingham, AL, USA

## Abstract

To date there have been no large multi ancestry genetic studies of osteoarthritis (OA). We leveraged the unique resources of 484,374 participants in the Million Veteran Program (MVP) and UK Biobank to address this gap. Analyses included participants of European, African, Asian and Hispanic descent. We discovered OA associated genetic variation in 10 loci and replicated association findings from previous OA studies. We also present evidence some OA-associated regions are robust to population ancestry. Drug repurposing analyses revealed enrichment of targets of several medication classes and provide potential insight to etiology of beneficial effects of antiepileptics on OA pain.

## INTRODUCTION

Osteoarthritis (OA) is a progressive joint disease with a poorly understood etiology. Disease management is limited primarily to symptom management (e.g., pain, inflammation). The societal and patient-centered impacts of OA among United States Veterans are profound with healthcare costs for treatment exceeding $880 million annually^1^. As no effective medical interventions for disease modification are available, OA often progresses to its end-stage at which time only surgical options are available, usually in the form of total joint replacement (TJR). A more thorough understanding of genetic influences of OA is essential to identify targeted personalized approaches to treatment, ideally long before end-stage is reached.

OA is a complex trait influenced by both genetic and non-genetic risk factors^2^. OA risk factors include aging, being female and/or overweight or obese^3^. However, OA is highly heritable with heritability estimates greater than 30%^4^. Although many genome-wide association studies (GWAS) have identified genetic loci associated with OA^5-12^, most studies have been performed in populations predominantly of European White descent including the largest GWAS in 826,690 subjects which identified 100 independent single nucleotide variants (SNVs) associated with OA across 11 OA phenotypes^13^.

To establish a more comprehensive map of genetic loci associated with OA, we performed multi-ancestry and ancestry stratified GWASes of 484,374 United States Veterans who participated in the Million Veteran Program (MVP) and participants from the UK Biobank (UKB). The combined cohorts comprise a larger number of participants from diverse ancestries and ethnicities. As such, we implemented analysis strategies to identify genetic variants associated with OA which may be study-specific, ancestry-specific or independent (**Figure 1**). We also performed transcriptome-wide imputation and fine-mapping to prioritize OA genetic signals as well as assessed heritability.

**Figure 1.**
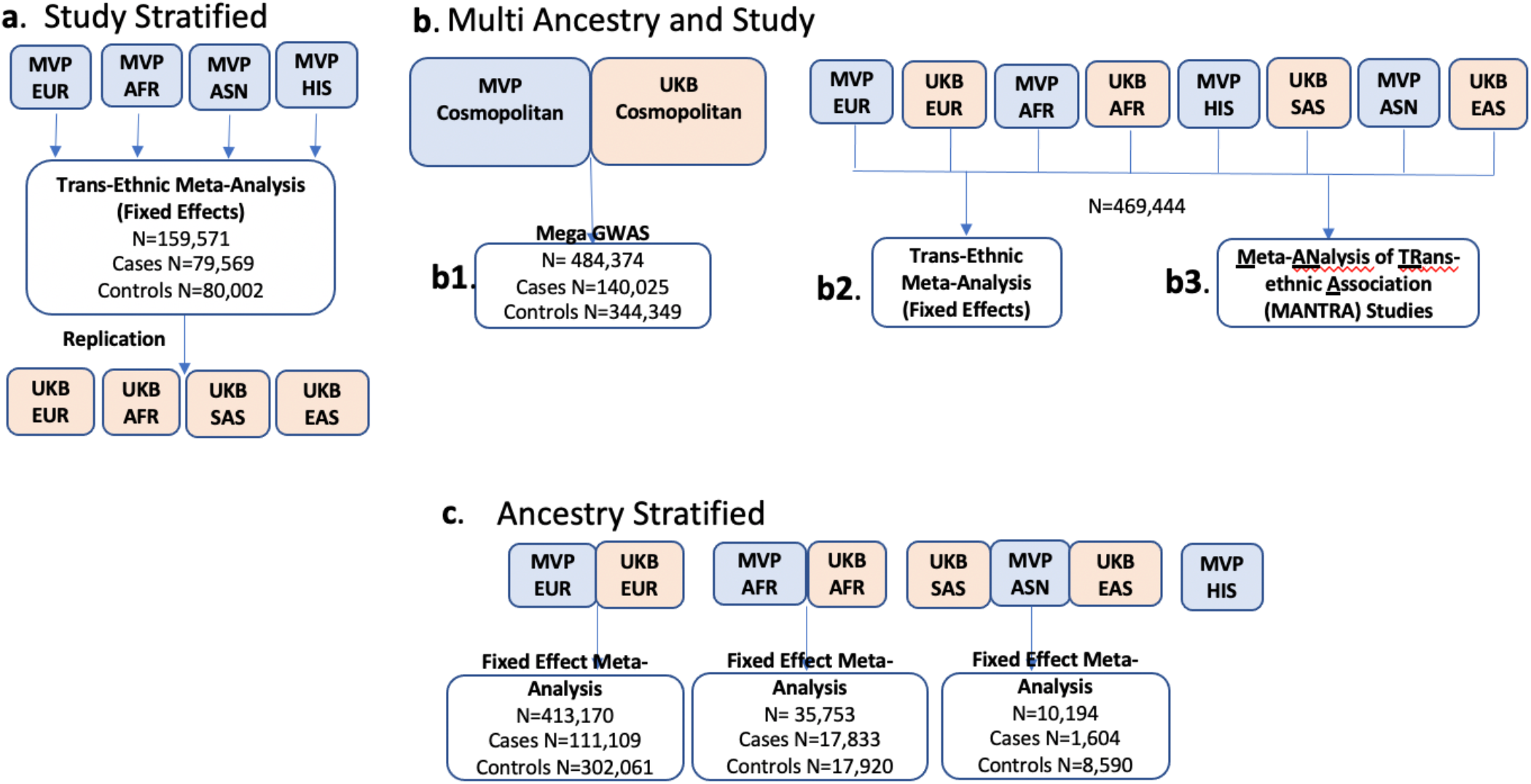
OA GWAS study design. **a. Study stratified** analyses with discovery genome-wide analyses were performed in MVP Veterans of European (EUR), African (AFR), Hispanic (HIS) and Asian (ASN) descent. Results were combined in a trans-ethnic meta-analysis and replication assessed in UKB EUR, AFR, South Asian (SAS) and East Asian (EAS) participants. **b. Multi Study and Ancestry** analyses for which three different approaches were used: **b1. Mega-analysis** of cosmopolitan MVP and UKB participants. No subject exclusion was made based on ancestry classification; **b2. Fixed effect meta-analysis**; and **b3. MANTRA** meta-analysis study and ancestry stratified analyses. **c. Ancestry stratified** analyses to identify specific genetic variants associated with OA, fixed effect meta-analyses were performed in EUR, AFR and ASN (SAS, EAS and MVP ASN) participants. GWAS was performed in HS Veterans however, no equivalent HIS replication group was present in UKB. Three analyses strategies were implemented to identify genetic variant which may be ancestry and study independent

## RESULTS

### Demographics of subjects included in analyses from MVP and UKB

In the MVP, 163,015 Veterans representative of four major racial/ ancestry groups were included in the analyses: 119,351 (73.2%) European White (EUR); 28,565 (17.5%) African American (AFR); 1,328 (0.8%) Asian (ASN); and 10,327 (6.3%) Hispanic (HIS) descent Veterans (**Table 1, Sup Figure 1)**. In the UKB, we also identified and included 321,359 subjects in analyses of four major ancestry groups (see HARE analysis in **supplementary materials** and **Sup Figure 2**): 293,819 (91.4%) EUR; 7,188 (2.2%) AFR; 1,704 (0.5%) East Asian (EAS); and 7,162 (2.2%) South Asian (SAS) (**Table 1**). In contrast to UKB, the MVP sample was predominantly male (**Table 1**).

**Table 1:** MVP and UKB descriptives.

|  | **MVP** |  |  |  |  | **UKB** |  |  |  |  |
| --- | --- | --- | --- | --- | --- | --- | --- | --- | --- | --- |
|  | *Cosmopolitan* | *European White* | *African American* | *Asian* | *Hispanic* | *Cosmopolitan* | *European White* | *African* | *East Asian* | *South Asian* |
| Number (% total) | 163,015 (100) | 119,351 (73.2) | 28,565 (17.5) | 1,328 (0.8) | 10,327 (6.3) | 321,359 (100) | 293,819 (91.4) | 7,188 (2.2) | 1,704 (0.5) | 7,162 (2.2) |
| Male % | 93.0 | 94.0 | 89.0 | 96.0 | 94.0 | 46.2 | 46.2 | 43.8 | 32.0 | 54.5 |
| Age (SD) | 63.4 (9.0) | 64.7 (8.6) | 59.8 (8.7) | 60.7 (10.2) | 60.9 (9.3) | 56.9 (8.1) | 57.1 (8.0) | 52.2 (8.0) | 52.8 (7.7) | 54 (8.5) |
| bmi (SD) | 30.0 (5.8) | 29.9 (5.8) | 30.4 (6) | 27.5 (4.4) | 30.6 (5.7) | 27.3 (4.8) | 27.2 (4.74) | 29.3 (5.3) | 23.9 (3.42) | 27.1 (4.4) |

MVP: Million Veterans Project. UKB: UK Biobank. OA: Osteoarthritis, Age: mean (standard deviation), bmi: mean (standard deviation), Cosmopolitan: Refers to entire available sample of participants without exclusions based on ancestry classifications.

**Figure 2:**
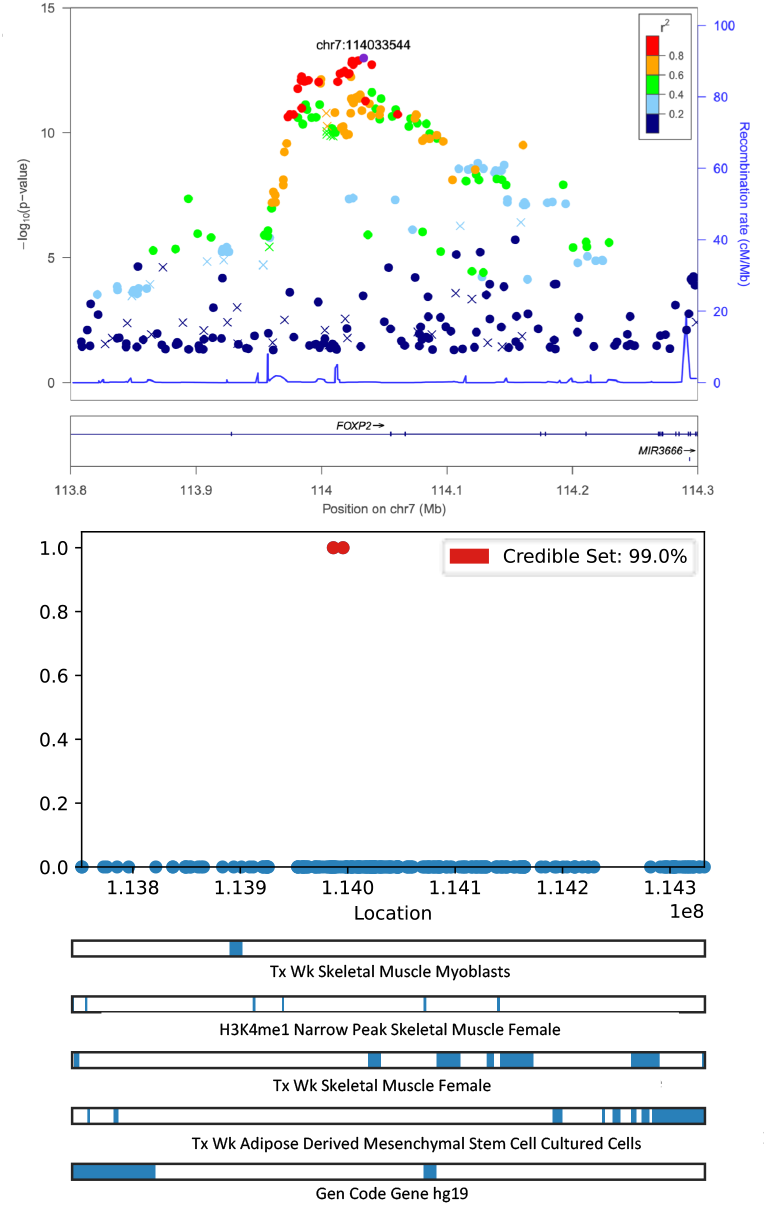
Fine mapping of FOXP2 region associated with OA in MVP trans-ethnic analysis: **Panel 1** Regional association plot; **Panel 2** functional annotation tracks. TxWk: Weak Transcription, H3K4me1: Histone 2 protein lysing 4 methylation, H3K36me2: Histone 3 protein lysine residue 36 methylation. Physical position denoted using hg19.

### Study stratified genetic variants associated with osteoarthritis

In the MVP, five genetic loci which we term MVP1 to MVP5 were significantly associated with OA in using fixed-effect multi-ancestry meta-analysis of 160,571 Veterans classified as either EUR, AFR, ASN or HIS descent (**Figure 1a**) and are summarized in **Table 2**. These loci included established and newly discovered OA associated loci: (1) *SLC39A8*^14^ on chromosome 4 (MVP1); (2) *FILIP1/SENP6/MYO6*^11,14,15^ on chromosome 6 (MVP2) and; (3) *DYNC1I1*^14,16^ on chromosome 7 (MVP3); (4) as well as a recently identified OA locus on chromosome 7 region encompassing *FOXP2*^*13*^ (MVP4); and (5) a 250 kb region on chr11q13 encompassing several genes including *KDM2A*^*13*^ (MVP5). In the MVP fixed effect meta-analysis, the p-values became smaller than that observed for the MVP EUR subset alone indicating contribution from non-EUR results to the significant meta-analysis results (**ST1**). Further, only MVP1, MVP3 and MVP4 loci had variants also nominally associated with OA in the UKB EUR GWAS **(ST1)**.

**Table 2:**
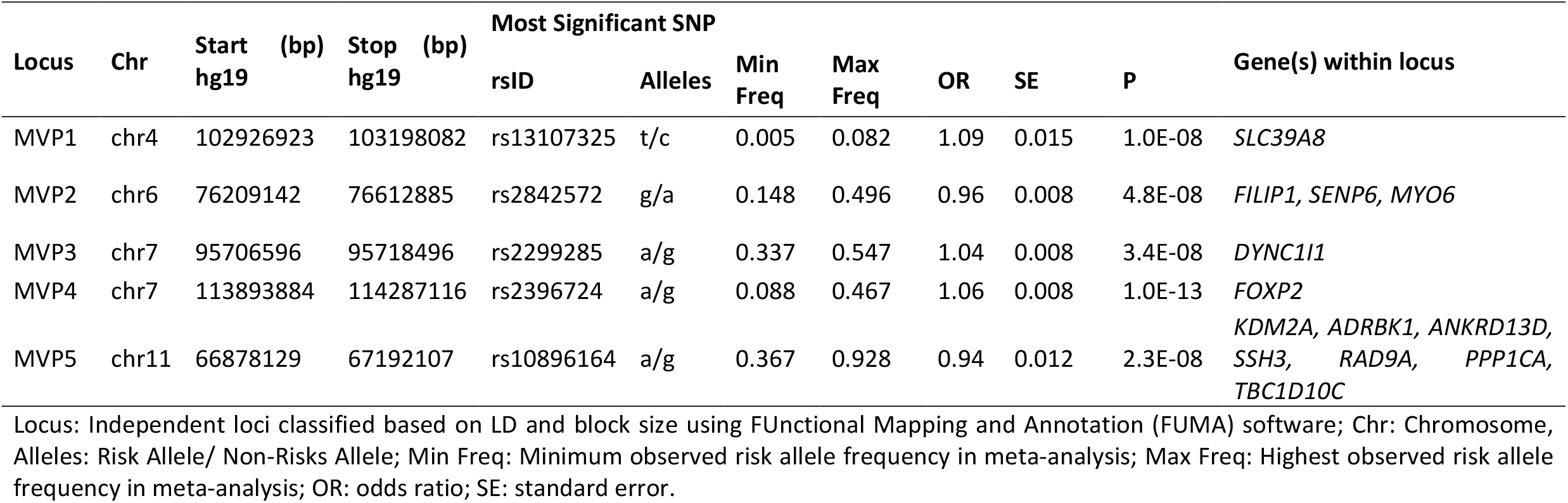
Genetic loci associated with OA reaching level of genome-wide significance (GWS) in the study stratified MVP GWAS.

A total of 138 genetic variants near and within the *FOXP2* gene were significantly associated (P<5.0E-8) with OA in the multi-ancestry meta-analysis of Veterans in MVP (**ST2**). Further, fine-mapping of the *FOXP2* region using PAINTOR refined the number of independent causal SNPs in the region to 2 SNPs, rs2049603 and rs1916980 (**Figure 2A**). These two SNPs are upstream in most *FOXP2* transcript variants and intronic in the two longest transcript variants, 7 (NR_033766.2) and X1 (XM_017012801.2). Both rs2049603 and rs1916980 variants replicated in the UKB EUR GWAS but not UKB AFR, SAS, or EAS stratified GWASes **(ST1)**. However, the direction of effect was consistent in UKB AFR and SAS **(ST1)**.

#### Multi-Ancestry OA Genetic Variants Associated with OA

As the biology relevant to a disease is not expected to differ across ethnic and/or ancestral groups, multi-ancestry results were primarily meta-analyzed using fixed effects meta-analysis. However, the frequency of causative OA genetic alleles and SNPs tagging causative alleles may differ across ancestries invalidating assumptions of fixed effect meta-analyses. For this reason, we employed and contrasted several approaches to generate multi-ancestry results (**Figure 1**). We used a linear mixed model (LMM) approach, implemented in BOLT-LMM^17^, to perform cosmopolitan GWASes in MVP and UK separately and then combined findings via fixed-effects meta-analysis which we refer to as the mega-GWAS (**Figure 1b**). Both the results of the mega-analyses and those of the cosmopolitan analyses in MVP and UKB are presented in **ST4**. We contrasted the results from the mega-GWAS with the two trans-ethnic meta-analyses strategies (**Figures 1b2 and 1b3**). A total of 27 genetic loci were significantly associated with OA in the MVP and UKB in the mega-GWAS (Table 3, **ST4, ST5, Figure 3**). Within the 27 loci, 476 SNPs were significantly associated with OA. Among these 476 SNPs, 50% were only significant using mega-GWAS analysis approach in contrast with fixed effect or MANTRA meta-analyses (**Figure 3a, ST6**). However, the majority of the 27 genetic loci were also discoverable using the fixed effect and MANTRA meta-analyses approaches (**Table 3**). Many of the loci encompass genes predicted to be significant eQTLs (**ST7**). Ten of the 27 loci in **Table 3** are novel not previously associated with OA. Among these loci, colocalization analyses indicated the credible set of variants may be influencing expression *PNPT1, EFEMP1, GORAPS, SCN11A, PTPRG, KDELR, DAGLB, CYP11B1, ARNTL, RIC8B, BTBD11* (**ST6**).

**Table 3:**
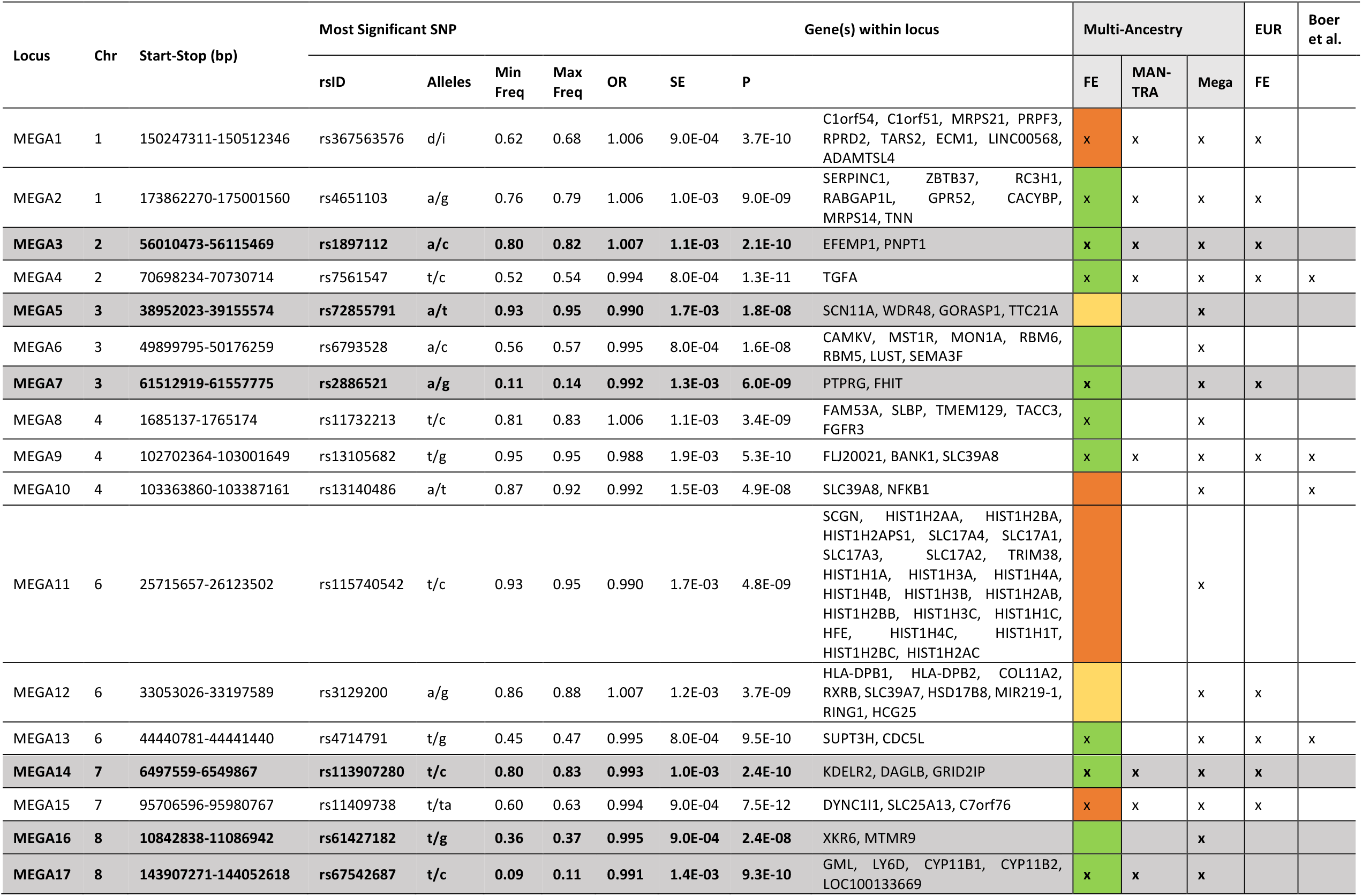
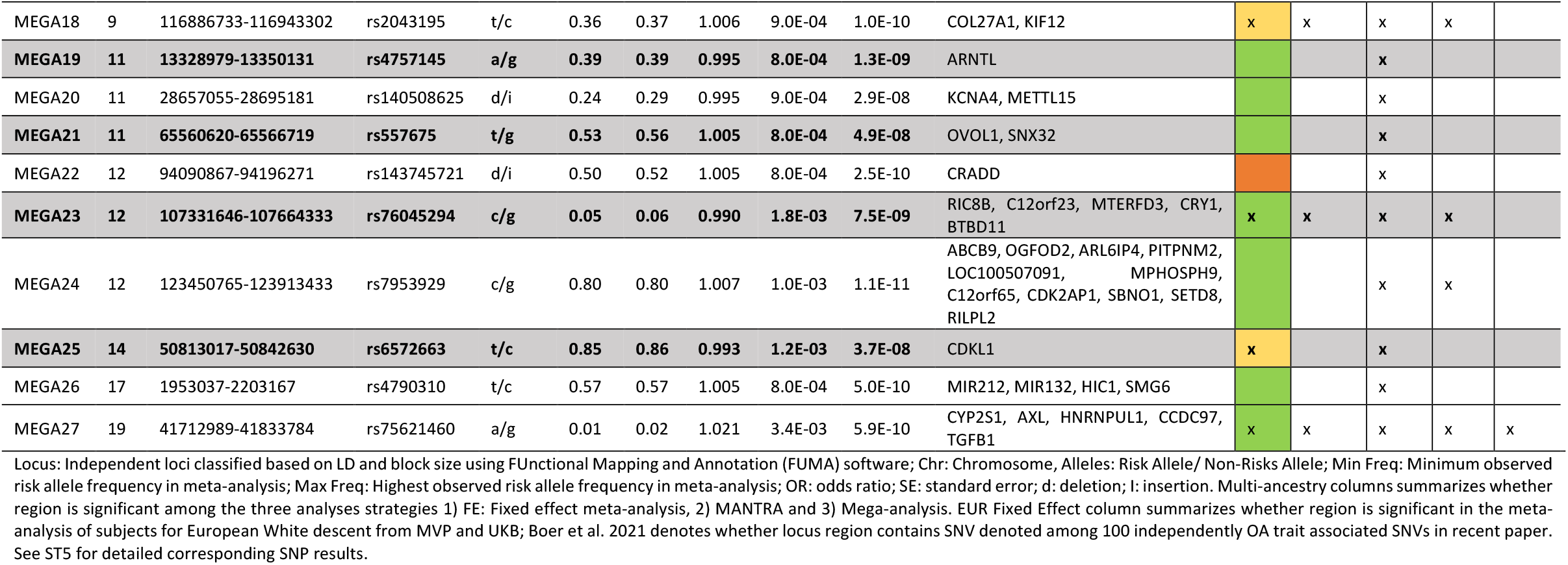
Genetic loci associated with OA reaching level of genome-wide significance (GWS) in the Mega GWAS analyses. Novel OA associated loci are highlighted in grey. Multi-ancestry fixed effect meta-analysis shading denotes highest I2 denoted for SNP in region where green I2<=30%, yellow 31-50% and orange 51-75%. No loci had I2>75%.

**Figure 3a:**
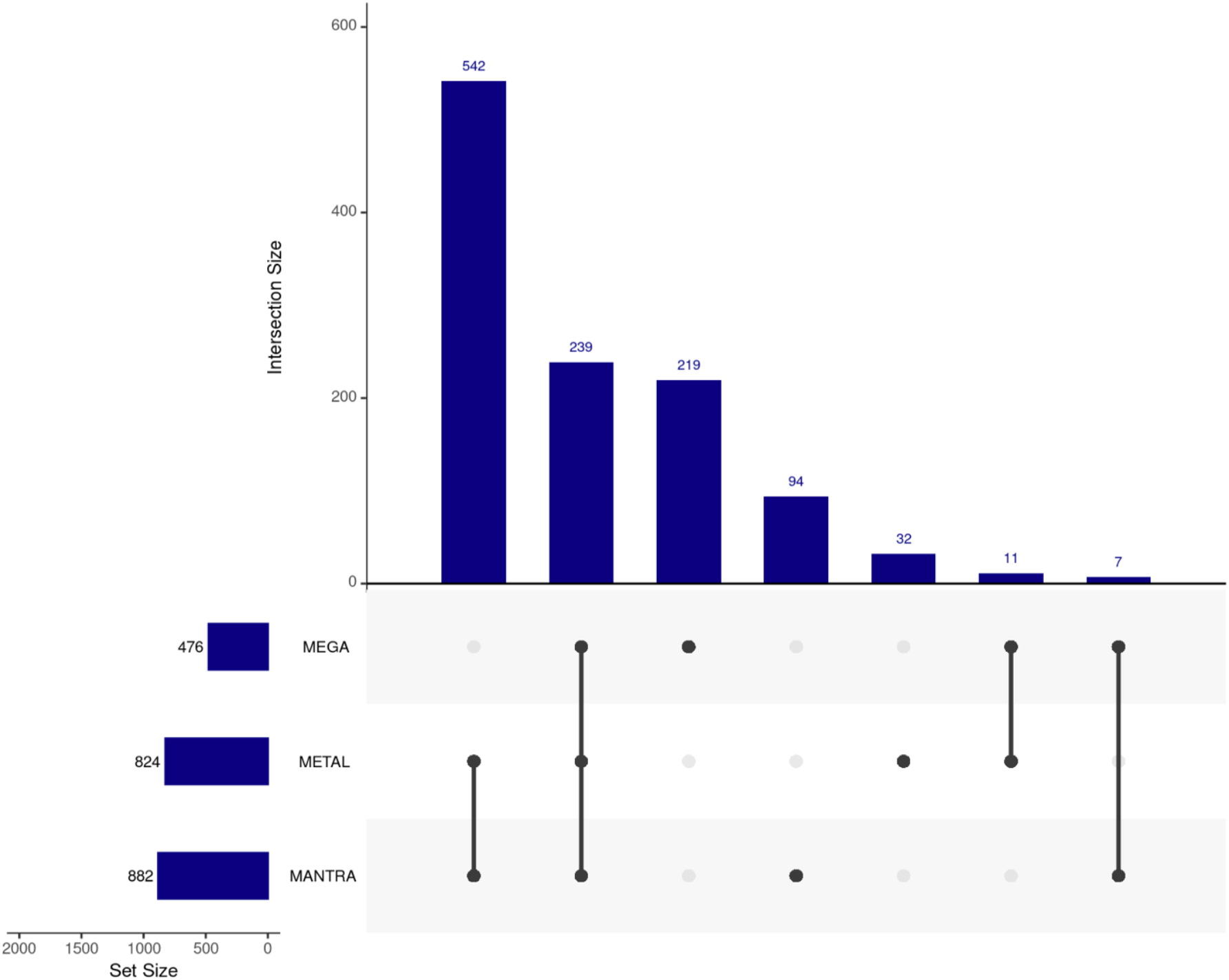
Upset plot comparing overlap and non-overlap of genetic variants signficiantly associated with OA mutli-ancestry analyses using either metal fixed effect, MANTRA trans-ethnic meta-analysis and mega analysis strategies outlined in Figure 1.

To gain insight to the contribution of allelic heterogeneity between the MVP and UKB ancestry groups, we examined heterogeneity of allelic effects (I^2^) from the fixed effect MVP+UKB in the context of the significant loci from the mega-GWAS (**Figure 3b**). No significant loci had SNPs that exhibited high heterogeneity (I^2^>75%) with most significant SNPs having low heterogeneity (I^2^<30%) in the MVP+UKB multi-ancestry fixed effect meta-analysis (**Table 3**). The correlation between effect sizes in MVP+UKB mega and fixed effect approached indicated the results were highly correlated R=0.96 (**Sup Figure 4**). A sensitivity analysis contrasting significant findings between fixed and random effects meta-analyses strategies did not reveal additional genetic variants associated with OA using random effects (**Sup Figure 5**).

**Figure 3b:**
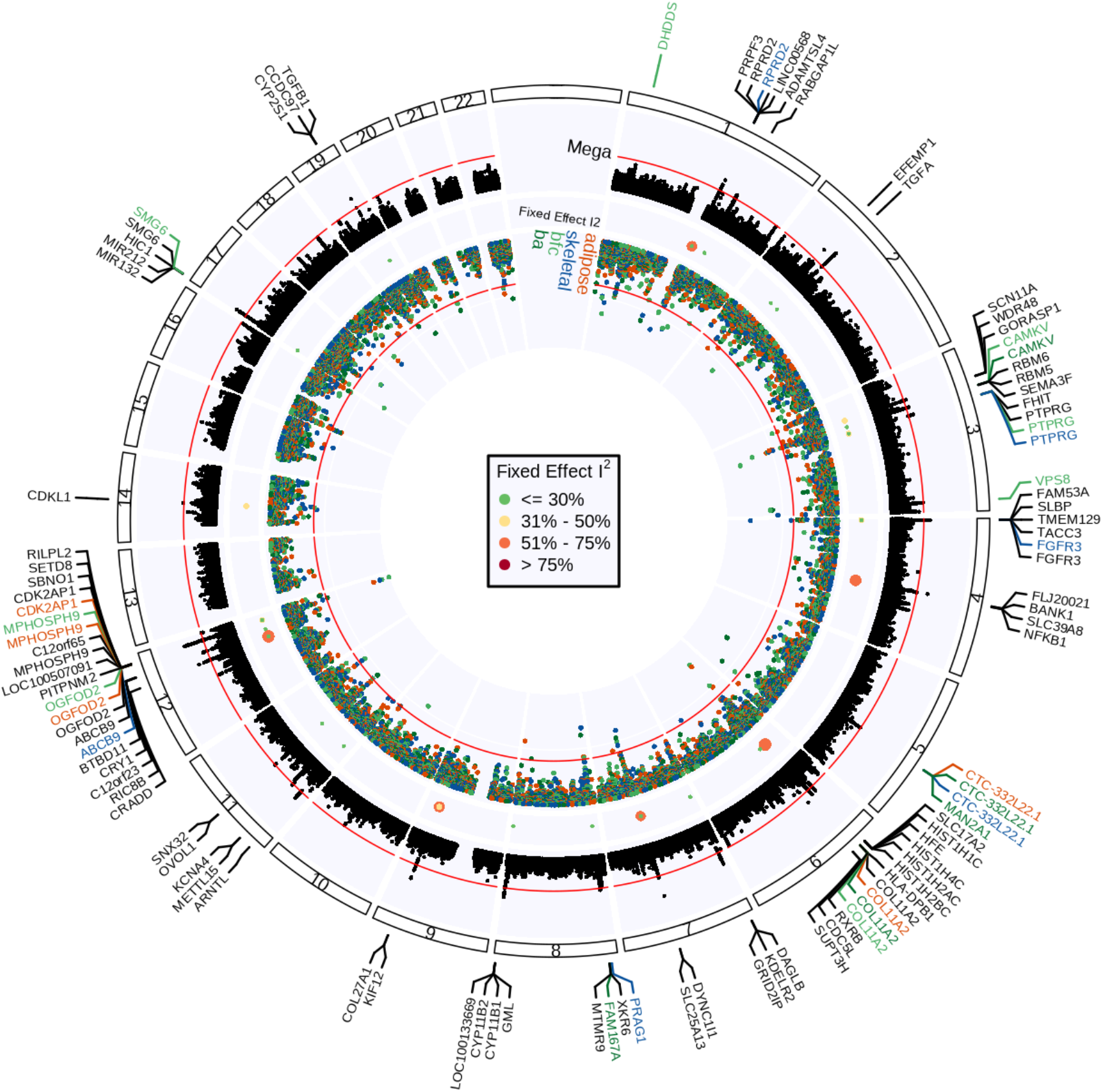
Summary of OA Mega-GWAS and transcriptome-wide imputation results. Outer ring displays -log10(P values) from the Mega-GWAS combining MVP and UKB results. GWAS significance is denoted at P=5E-8 by red line. Middle ring denotes the fixed effect I2 from the meta-analysis of MVP: EUR, AFR, ASN and HIS and UKB: EUR, AFR, SASN, EASN. Innter track denotes display -log10(P values) of corresponding expression quantitative trait loci (eQTL) in **adipose, skeletal, brain amygdala (ba)**, and **brain frontal cortex (bfa)**, respectively. eQTL significance is denoted at P=1E-5 by red line.

### Ancestry stratified genetic variants associated with OA

Ancestry stratified analyses were used to highlight genetic variants which may be ancestry independent in EUR, AFR, ASN and HIS populations (**Figure 1c**). Among EUR MVP+UKB participants, 32 independent loci (**ST9**) were significantly associated with OA (**ST9, ST10**). Many of the OA associated genetic loci were predicted to contain eQTLs in adipose, skeletal muscle, brain frontal cortex and/or brain amygdala tissues (**ST11**). The most significant *FOXP2* SNP from the MVP multi-ancestry analysis, rs2396724, remained significantly associated in MVP-EUR but was only nominally significant in UKB-EUR. In the fixed effect meta-analysis of MVP+UKB EUR, there was suggestion of heterogeneity of effects for rs2396724 (Phet < 0.10).

Among Veterans of AFR descent, variants in three regions (*MYO5B; CCDC11*), (*LINC00314-LINC00161*), and (*MCTP2-LOC440311*) were associated (P<5E-8) with OA (**ST12**). In the MVP+UKB AFR meta-analysis, only SNPs in *LINC00314-LINC00161* region variants achieved GWS (e.g., rs76386041, MAF=0.03, OR=0.76, SE=0.05, P=1.6E-8); however, this result was less significant than the finding in the MVP AFR GWAS alone (e.g., rs76386041, MAF=0.033, OR=0.75, SE=0.05, P=1.1E-8). In the transcriptome wide imputation, expression of the *MYO5B* gene in adipose tissue was predicted to be significantly associated with OA (**ST13**).

No variant was associated with OA at a level of GWS among ASN Veterans. The most significant variants associated with OA among ASN Veterans are intronic to the *RELN* (rs147959272, MAF=0.12, OR=2.4, SE=0.17, P=2.4E-7) and *MYLK* (rs11409032, MAF=0.26, OR=2.0, SE=0.14, P=7.7E-7) genes (**ST14**).

Among Veterans of HIS descent, one locus, encompassing the gene *GUCY1B3*, had several variants associated with OA at level of GWS (e.g rs58196867, MAF=0.02, OR=0.44, SE=0.17, P=2.8E-9, **ST15**). The *GUCY1B3* gene encodes soluble guanylate cyclase, the expression of which is downregulated in cartilage precursor cells under high hydrostatic pressure^18^. Unfortunately, our HARE analyses did not reveal a corresponding Hispanic group in the UKB (**Supplementary Methods**). For this reason, confirmation of our findings among Veterans of HIS descent were assessed in all available UKB ancestry groups (EUR, AFR, EAS, and SAS). No evidence for replication of the association between the locus and OA was found in these groups **(ST15)**.

#### Association of literature reported OA variants

We examined whether previously reported OA variants^12^ were also associated with OA in the current findings. A total of 72 (64%) SNPs were found to overlap in current findings with consistent direction of association and nominal significance (**ST16**). SNPs within *COL27A1, DYNC1L1* and *GSDMC* gene regions were also associate in both MVP EUR and AFR ancestry groups as well as in the MVP+UKB MANTRA analysis. SNPs within *ANAPC4, FAM53, PPP1R3B*, and *TGFA* gene regions were also associated with OA in both the MVP EUR as well as with MVP+UKB MANTRA. One variant within *HDAC9*, rs11764536, was also associated in the MVP HIS as well as in the MVP+UKB MANTRA analysis. Several variants were also associated only in a single MVP ancestry group such as variants within *BMP*5 (MVP AFR), *RUNX2* (MVP AFR), *SLC44A2* (MVP ASN) and *LMX1B* (MVP AFR). Further several SNPs were associated only in the MVP EUR including variants within *CHADL, GNL3/ITIH, ITIH1* and *ITIH5* gene regions.

One SNP within the *SLC44A2* region was also associated in MVP ASN. A previously reported OA SNP, rs7222178, within the *NACA2* gene region was nominally significant in MVP ASN with seemingly opposite direction of effect. However, rs7222178 is an A/T variant which is difficult to assess uniformity in direction of effect due to known strand problems with A/T and G/C variants. We then also noted other AT/GC variants which appear to be also associated in terms of effect direction and p-value: *ANAPC4, COLD27A1, DPEP1* and *ITIH1*. Several variants were also associated only in the combined MVP+UKB MANTRA analyses including those within *DPEP1, FGF18, GDF5, GLIS3, HLA-DP1, LTBP1, LYPLAL1-AS1, RAB28, RBM6* and *TGFB1* gene regions.

#### Gene-set enrichment analyses of genes with SNPs associated with OA

To gain insight to the overall etiological role of genetic variation in OA, we examined enrichment of genes with SNPs identified as associated with OA in the current discovery and/or replication analyses (**ST16** and **ST17**). Gene-set enrichment analyses enrichment of genes from 616 gene-sets among which are many are known to contribute to the etiology of OA including cartilage development, chondrocyte differentiation, connective tissue development, skeletal system development, ossification, bone mineralization, bone morphogenesis and elastic fibers. Both newly discovered and previously associated OA genes were represented in these OA pathways (**ST18)**. Additionally, many OA genes were members of biosynthetic process pathways including cortisol, mineralocorticoid, glucocorticoid, aldehyde and phosphorus.

#### Heritability of osteoarthritis

Heritability of OA was estimated in MVP and UKB populations with at least 10,000 subjects (MVP: EUR, AFR, HIS and UKB: EUR, **Table 4**). In MVP, heritability of OA was highest among Veterans of Hispanic descent (51.3%). OA heritability was lowest for participants of EUR descent at 19.8% and 8.2% in MVP and UKB, respectively. The proportion of heritability in each population accounted for variants identified as eQTLs in our MetaXcan analyses was low with the highest estimate being 0.89% in the MVP EUR.

**Table 4.** Narrow sense heritability of osteoarthritis in MVP and UKB cohorts estimated using BLD-LDAK.

| Ancestry | MVP % $h^2$ (SD) | | | UKB % $h^2$ (SD) | | |
| --- | --- | --- | --- | --- | --- | --- |
|  | N | Total | OA eQTL | N | Total | OA eQTL |
| EUR | 119,351 | 19.8 (1.4) | 0.89 (0.10) | 293,819 | 8.2 (0.83) | 0.67 (0.070) |
| AFR | 28,565 | 26.1 (5.7) | 0.56 (0.17) | -- | -- | -- |
| HIS | 10,327 | 51.3 (8.7) | 0.38 (0.26) | -- | -- | -- |

**Abbreviations:** AFR – African; EUR – European; $h^2$ – narrow sense heritability; HIS – Hispanic; MVP – Million Veterans Program; UKB – UK Biobank; SD – Standard Deviation; -- sample size less than 10,000.

#### Potential for drug repurposing

We examined whether genes predicted to be OA eQTLs using MetaXcan were enriched for targets of currently approved medication classes. Using the set of predicted eQTLs from the MVP+UKB mega-GWAS, statistically significant enrichment of targets for three classes of drugs was observed: 1) anti-neoplasmic agents (OR: 1.7, Pexact = 0.014); 2) anti-epileptics (OR: 2.0, Pexact = 0.026); and 3) anti-acne preparations (OR: 3.2, Pexact = 0.014, **ST19**).

## Discussion

We presented results of a multi-ancestry GWAS of OA in 484,374 MVP Veterans and UKB participants. In doing so, we discovered both ancestry specific genetic variants associated with OA as well as genetic variants which may be robust to genetic ancestry. OA biology is not expected to differ across ethnic and/or ancestral groups. Race and ethnicity are social constructs which have contributed to discovery bias in human genomics^19^. In the ideal setting, genetic risk alleles contributing to OA etiology are uniformly present and penetrant in all ancestry populations investigated. However, the frequency of causative OA genetic alleles and SNPs tagging causative alleles may differ across ancestries invalidating assumptions of fixed effect meta-analyses for this reason we used several to generate multi-ancestry results. Although we performed multi-ancestry and stratified analyses, most subjects in the MVP and UKB studies are of European White descent. There remains a great need for more research to identify genetic variants and regions associated with OA which are robust to ancestry for developing precision medicine approaches generalizable to all patients. Our multi-ancestry genetic association study of OA represents only a step towards closing the gap in the lack of studies investigating the genetics of OA in populations of non-European White descent.

More specifically, we identified 10 novel loci associated with OA using the multi-ancestry mega-analysis approach. These loci included SNPs on chromosome 2 within the *EFEMP1* gene encoding the protein fibulin-3. Fibulins are known to contribute to elasticity and functional integrity of connective tissues^20^. As a biomarker, fibulin-3 level in serum and urine correlate with OA disease incidence and progression^21-23^. Interestingly, fibulin-3 has been used to monitor efficacy in randomized control trials of OA treatments^22-24^. Additionally, SNPs within and intergenic to the genes *SCN11A, WDR48, GORASP1* and *TTC21a* on chromosome 3 were associated with OA. Among these, *WDR48* encodes a protein influencing ubiquitination of proteins including *PHLPP1* whose expression promotes OA progression^25^. Further, *SCN11A*, is known to play a role pain perception^26^. At a second locus on chromosome 3, SNPs within the gene *PTPRG* were associated with OA. *PTPRG* encodes the protein tyrosine phosphatase, receptor type, G which has been shown to be hydryoxymethylated in chondrocyte tissue from patients with OA^27^. On chromosome 7, SNPs within a novel region encompassing the genes *KDELR2* and *GRID2IP* were associated with OA. Colocalization results implicated expression of *KDELR2* in skeletal muscle at this locus (**ST6**). Genetic mutations in *KDELR2* cause severe, progressively deforming osteogenesis imperfecta^28^. Independent loci on chromosomes 11 and 12 were associated with OA that contained genes (*ARNTL* and *CRY1*) known to regulate the circadian clock^29^ which has been implicated in osteoarthritis^30^.

Despite several large GWAS studies of OA among participants of EUR descent including the recently published de Boer et al 2021^13^, we still identified 4 novel loci significantly associated with OA in the EUR stratified results from MVP and UKB. These include: 1) EUR3: a region on chromosome 2 encompassing the gene *EFEMP1*; 2) EUR4: a region on chromosome 3 encompassing the genes *SCN11A, WDR48, GORASP1* and *TTC21A*; 3) EUR19: a region on chromosome 8 encompassing the genes *GML, CYP11B1* and *CYP11B2*; and 4) EUR26: a region on chromosome 12 encompassing the genes *TMEM263, MTERF2 and CRY1*. These 4 novel loci were also significantly associated with OA in the mega-analyses combining the cosmopolitan analyses in MVP and UKB discussed above. This may indicate that although these loci were discoverable in EUR populations there is evidence the association with OA may be ancestry independent.

In the current report, we summarized the results of several strategies for analyzing multi-ancestry and study data. The mega-analysis approach discovered 10 independent loci associated with OA. These include 4 novel OA associated loci (MEGA5, MEGA16, MEGA19 and MEGA21) not GWS significant in the fixed effect multi-ancestry or fixed effect meta-analysis results. For all 4 loci (see **ST5**) the corresponding P-values for lead SNPs were at near GWS using the fixed effect multi-ethnic and EUR specific analysis approach. However, we believe the increase in power enabling identification of these four loci occurred was for several reasons. Stratification and combining results by meta-analysis is expected to have lower power than mega-analysis^31,32^. Thus, in the current study the mega-analysis approach was more powerful due to both increased sample size and lack of stratification of the cohort. Second, we employed an LMM approach implemented in BOLT-LMM which is designed to control for both population structure and relatedness. LMM models have been shown to be robust for controlling for population structure and relatedness. Finally, the practice of analyzing a complete cohort and/ or pooling studies via mega-analysis is not novel^31,32^. This approach is increasingly used to identify variants associated with disease from whole-genome sequencing. These include several publications from the Trans Omics for Precision Medicine (TOPMed) consortium where both stratified and pooled ancestry/ study analyses are presented with the latter described as most powerful^33-36^. A large concern for all of the meta-analysis strategies employed in the current manuscript was that more than 75% of participants analyzed from both MVP and UKB are of European White descent which may mask important signals frequent only in the African, Hispanic and Asian descent groups. For this reason, we also performed ancestry stratified analyses.

Further, we identified several genetic regions associated with OA among African American, Hispanic and Asian descent Veterans. In our analysis of 29,225 African American Veterans, *MYO5B* was both significantly associated with OA and predicted to be differentially expressed in adipose tissue. Genetic variation in *MYO5B* has been previously associated with HDL cholesterol^37^. Additionally, observations of elevated cholesterol in synovial fluid from OA patients have led to the hypothesis cholesterol metabolism may contribute to OA pathogenesis^38^. Further research into *MYO5B*’s role in OA pathogenesis via cholesterol metabolism is merited as hypercholesterolemia can be monitored and treatments exists for controlling it. We also found evidence for replication of many previously associated OA genetic variants including some replicating among AFR, HIS and ASN descent Veterans. This may indicate some of the genetic variants and regions associated with OA in previous, largely European White populations are robust to genetic ancestry.

Overall, we identified 31 genetic loci associated with OA in the mega analysis. Among these, 7 loci were only discoverable as OA associated loci in MVP+UKB multi-ancestry analyses. Discovery of these 7 OA associated genetic regions in MVP+UKB multi-ancestry analyses provides support findings are not exclusively driven by the large sample size of the combined MVP+UKB. However, some previously reported OA associated loci did not replicate. The frequency of causative OA genetic alleles and SNPs tagging causative alleles may differ across ancestries. As the majority of previous GWAS in OA begin performed in populations of European descent it is challenging to disentangle whether lack of replication in a diverse population such as MVP implies the locus is EUR specific, not relevant to our understanding of OA and/ or limited by small sample size of diverse ancestral groups. We used several state-of-the-art approaches for GWAS in multi ancestry cohorts however, demonstrate the majority of the loci were discoverable in the EUR stratified analyses (**Table 3**). Although the mega-analysis had the largest sample size and identified more SNPs significantly associated with OA, the EUR stratified approach identified slightly more independent loci (**ST8**). However, several of these loci were not significant in the mega-analysis indicating these loci may be largely EUR specific. As populations become increasingly more admixed previously associated OA EUR loci are likely to persist in terms of frequency. Thus, there remains a need to refine previously associated OA loci by developing new techniques for fine mapping and analysis methods robust to analyses of multi ancestry populations. This is in addition to increasing the sample size of non-EUR populations investigated.

Further, gaining biological insight from the identified genetic associations is complicated by the presence of multiple trait (pleiotropic) associations within the same locus. For example, we identified a novel 250 kb region on chr11q13 encompassing several genes associated with OA in MVP and previously associated with total hip replacement^12,13^. Interestingly, this locus has also been associated with body fat distribution^39^ and bone mineral density^40^ among other traits like body height^41-43^ and lipid levels^44^. Being overweight and having high bone mineral density are established risk factors for OA with evidence supporting a direct effect of bone density on joint deterioration^45^ and BMI^46^ on osteoarthritis. Similarly, variants in 11p15.3, one of the other identified novel loci (MEGA19), is associated with height^47^, BMI^42,48^, lipids^49^, blood pressure^50^ and CRP levels^51,52^. Such pleiotropic relationships are pervasive across complex traits^53-56^ make it very difficult to pinpointing the pathways mediating the identified associations with risk of OA.

Interestingly, our drug repurposing findings provide support for current physician’s intuition in terms of prescription patterns of anti-epileptics for OA pain. The anti-epileptics gabapentin and pregabalin are increasingly prescribed to patients with OA to help with pain despite gaps in understanding how they work for OA patients^57^. We further discovered OA eQTLs were significantly enriched with targets of anti-neoplastic agents and anti-acne preparations. Many anti-neoplastic agents have anti-fibrotic properties^58,59^. Synovial inflammation and fibrosis mark OA progression^60^ supporting logic some anti-neoplastic agents may have benefits for OA. Anti-acne preparations included azaleic acid which is decreased in urine in a OA rat model^61^. Additional research is merited to test whether azaleic acid in urine could serve as biomarker of OA progression in humans. Further, beta-blocker use has been associated with less OA joint pain^62^. The G Protein-Coupled Receptor Kinase 2 (GRK2), encoded by the gene *ADRBK1*, has been shown to regulate beta-adrenergic receptor (β-AR) expression and downstream activity influencing sensitivity to pain^63,64^. In our study stratified analyses in MVP, rs10896164 variant, intronic to *ADRBK1* was significantly associated with OA in MVP EUR, AFR, HIS and ASN ancestry Veterans. Thus, several OA associated loci provide etiological foundation for repurposing of approved drugs.

Gene-set enrichment analyses of genes with SNPs previously associated with OA as well as newly discovered SNPs within or near novel genes highlighted OA relevant gene-sets. For example, the rs532464664 variant frameshift mutation in the *CHADL* gene was previously identified as a high risk for hip OA variant in the Icelandic population^65^. *CHADL* is expressed in cartilage and the frameshift mutation leads to decay of the mutant transcripts^65^. In the current analyses, the *CHADL* frameshift mutation was rare (MAF=0.05%) replicating in MVP EUR with a large effect size but not MVP AFR, HIS or ASN. Furthermore, several of the newly OA associated genetic variants are in or near genes belonging to the same gene-sets as *CHADL* (**ST19)**. Enrichment of novel OA associated loci genes along with *CHADL* in cartilage, chondrocyte and skeletal muscle development and differentiation gene-sets strengthens the support for their role in OA.

Our study has strengths and limitations. Analyses to discover SNPs associated with OA were performed in, to our knowledge, the largest multi-ancestry cohort to date. Another strength is the implementation of methodology to identify non-EUR descent participants in UKB. Most research using the UKB has focused on subjects of self-identified EUR descent. To address this limitation, we used the HARE^66^ approach to classify UKB subjects of EUR, AFR, SAS and EAS descent. The HARE approach is an expansion of methods that use genetic data to characterize race by also harmonizing self-reported race and ethnicity in the classification algorithm. In doing so, additional insight was gained as to whether previously identified OA genetic regions were associated in populations of non-EUR descent. Identification of a panel of genetic variants associated with OA independent of ancestry would enable the development of an OA polygenic risk score robust to population structure and assist in the development of personalized medicine approaches to OA. We also followed up findings using fine mapping, transcriptome-wide imputation analyses and drug repurposing to prioritize genes from signals where multiple genes and variants were significantly associated.

Study limitations include the lack of radiographs that could potentially improve the validity of the electronic health record (EHR) diagnostic codes used to code OA. We mitigated this limitation somewhat by coding OA in MVP only if a Veteran had two OA codes more than 30 days apart as well as attempting to replicate significant findings in the UKB cohort. To our knowledge, this is the first analysis of OA coded from EHR data to require two OA codes more than 30 days apart. Ideally, our analyses would have included a sensitivity analysis to determine if findings are robust when OA status is confirmed by radiograph gradings. Nonetheless, the frequency of OA was higher in MVP than UKB. Being older in age and having higher BMI are the strongest risk factors for OA^67,68^. Although MVP Veterans are predominantly male (lower risk), in keeping with the strongest risk factors MVP Veterans were both slightly older with higher BMI than UKB participants. This is keeping with previous reports of OA being higher among Veterans than non-Veterans^69-71^. Although we excluded Veterans with post-traumatic OA, Veterans overall have a higher risk of physical trauma, an occupational hazard, which is also a strong risk factor for OA^68,72^. Given our stringent exclusion criteria, many MVP participants were excluded from the control population, which could hypothetically introduce bias by creating a population of super controls. However, the allele frequencies of our control populations in the MVP cohort closely match relevant 1000 Genomes Project reference superpopulations making it unlikely that our exclusion criteria were a source of significant bias. Although this was the largest multi-ancestry genetic analysis of OA to date, sample sizes for Veterans of ASN descent were small and replication of our findings for the HIS descent Veterans was not possible in the UKB cohort. This limitation calls for additional research in larger multi-ancestry studies with higher numbers of participants of Asian and Hispanic descent. Finally, a possible limitation to the analyses is the traditional P<5×10^−8^ was employed as the level of genome-wide significance in the analyses. The body of research on appropriate GWS thresholds in more diverse studies analyzing rarer variants is evolving^73-76^. However, in the current study additional validation was provided thru the observation many of the newly associated OA loci encompassed variants annotating to genes in gene-sets known to play a role in etiology of osteoarthritis.

In sum, we have identified new and replicated known genetic regions associated with OA. Multi-ancestry associated OA variants identified in the current report only begin to fill the void in identification of ancestry independent OA genetic variants needed to advance the development of polygenic risk scores. Larger sample sizes of ancestrally and ethnically diverse subjects are needed to continue to fuel progress and remedy discovery bias imposed by previous EUR focused research.

## Data Availability

Summary Statistics will be made available in dbGaP phs001672.v7.p1

---

**Figure 1: OA GWAS study design: a) Study stratified** analyses with discovery genome-wide analyses were performed in MVP Veterans of European (EUR), African (AFR), Hispanic (HIS) and Asian (ASN) descent. Results were combined in a trans-ethnic meta-analysis and replication assessed in UKB EUR, AFR, South Asian (SAS) and East Asian (EAS) participants. **b) Multi Study** analyses for which three different approaches were used: **b1) Mega-analysis** of cosmopolitan MVP and UKB participants. No subject exclusion was made based on ancestry classification; **b2) Fixed effect meta-analysis**; and **b3) MANTRA** meta-analysis study and ancestry stratified analyses. **c) Ancestry stratified** analyses to identify specific genetic variants associated with OA, fixed effect meta-analyses were performed in EUR, AFR and ASN (SAS, EAS and MVP ASN) participants. GWAS was performed in HIS Veterans however, no equivalent HIS replication group was present in UKB.

**Figure 2: Fine mapping of FOXP2 region associated with OA in MVP trans-ethnic analysis**: **Panel 1** Regional association plot; **Panel 2** functional annotation tracks. TxWk: Weak Transcription, H3K4me1: Histone 2 protein lysing 4 methylation, H3K36me2: Histone 3 protein lysine residue 36 methylation. Physical position denoted using hg19.

**Figure 3: Summary of OA Mega-GWAS and transcriptome-wide imputation results**. Outer ring displays -log10(P values) from the Mega-GWAS combining MVP and UKB results. GWAS significance is denoted at P=5E-8 by red line. Middle ring denotes the fixed effect I2 from the meta-analysis of MVP: EUR, AFR, ASN and HIS and UKB: EUR, AFR, SASN, EASN. Innter track denotes display -log10(P values) of corresponding expression quantitative trait loci (eQTL) in **adipose, skeletal, brain amygdala (ba)**, and **brain frontal cortex (bfa)**, respectively. eQTL significance is denoted at P=1E-5 by red line.

**Supplementary Table 1:** Independent significant SNPs associated with OA in MVP at level of genome-wide significance (GWS) and corresponding results in UKB ancestry groups.

**Supplementary Table 2:** Multi-Ancestry genetic variants significantly (p<5×10^−8^) associated with osteoarthritis in the Million Veteran Program (MVP) using fixed effects meta-analysis.

**Supplementary Table 3:** Differentially Expressed Adipose, Brain, and Skeletal Muscle Genes Associated with Osteoarthritis in MVP Veterans Using Transcriptome-wide Imputation

**Supplementary Table 4:** Mega-analysis genetic variants significantly associated with osteoarthritis in MVP and UKB.

**Supplementary Table 5:** Mega-analysis independent genetic loci significantly associated with osteoarthritis in MVP and UKB.

**Supplementary Table 6:** Genetic loci associated with OA reaching level of genome-wide significance (GWAS) in Meta-analysis Million Veteran Program (MVP) and UK Biobank (UKB) participants of European descent (EUR).

**Supplementary Table 7:** Genetic variants associated with osteoarthritis in MVP and UKB using trans-ethnic (MANTRA) and fixed effect meta-analyses.

**Supplementary Table 8:** MVP and UKB transcriptome-wide imputation of mega-analysis GWAS of osteoarthritis results. Mega-GWAS results were input to MetaXcan to impute transcriptome using GTEx version for 4 tissues, adipose, brain amygdala, brain frontal cortex, and skeletal muscle.

**Supplementary Table 9:** Genetic loci associated with OA reaching level of genome-wide significance (GWS) in Meta-analysis Million Veteran Program (MVP) and UK Biobank (UKB) participants of European descent (EUR).

**Supplementary Table 10**: Genetic variants significantly associated with osteoarthritis in Meta-analysis Million Veteran Program (MVP) and UK Biobank (UKB) participants of European descent (EUR).

**Supplementary Table 11:** MVP and UKB European descent participants transcriptome-wide imputation of fixed effect osteoarthritis meta-analyses results.

**Supplementary Table 12:** Genetic Variants Associated (p < 1×10^−4^) with Osteoarthritis in Meta-Analysis of Million Veteran Program (MVP) and UK Biobank (UKB) participants of African Descent (AFR).

**Supplementary Table 13:** MVP and UKB African descent participants transcriptome-wide imputation of fixed effect osteoarthritis meta-analyses results.

**Supplementary Table 14**: Top 10 meta-analysis genetic variants associated with osteoarthritis in Million Veteran Program (MVP) and UK Biobank (UKB) participants of Asian descent.

**Supplementary Table 15**: Top 10 genetic variants associated with osteoarthritis in Hispanic descent Veterans from the Million Veteran Program (MVP).

**Supplementary Table 16:** Replication status of variants previously associated with OA.

**Supplementary Table 17**: Summary of genes with genetic variants associated with OA in current analyses (both discovery and replication).

**Supplementary Table 18**: Gene-set enrichment analyses of genes associated with OA in current analyses (both discovery and replication).

**Supplementary Table 19**: Evidence supporting drug repurposing for OA based on genes predicted to be significantly (P<0.05) in transcriptome-wide imputation of mega-GWAS results from MVP and UKB.

**Supplementary Table 20**: ICD9 and ICD10 inclusion codes used to code osteoarthritis.

**Supplementary Table 21**: Exclusion codes applied to non-osteoarthritis defined controls in MVP and UK Biobank

## METHODS

### Ethical approval

The work described in this manuscript received ethical and study protocol approval from the Veterans Affairs Central Institutional Review Board as well as the University of Alabama at Birmingham in accordance with the principles outlined in the Declaration of Helsinki. UK Biobank data was analyzed under approved access via proposal #30350.

### Osteoarthritis trait definition

Osteoarthritis cases and controls were identified using International Classification of Disease, 9^th^ revision, common modification (ICD-9-CM) and ICD-10-CM codes from the MVP diagnosis file version 18.2. Diagnosis dates ranged from July 2, 1990 to June 13, 2019. ICD code lists from Zengini et al. ^77^ were used. For OA case identification, lists of codes were provided for a general OA category and overlapping lists for specific subtypes of OA (Hip, Knee, Spine, Hand, Finger, and Thumb) provided in **Supplementary Table 20**. OA cases were not restricted to any OA subtype(s) and included OA cases with any hip, knee, hand, finger, thumb, spine and other site OA ICD codes (**Sup Figure 1**). The exclusion criteria for controls included codes for frequent concomitant findings typically more prevalent in patients with OA such as chondrocalcinosis, crystal arthropathies, bony abnormalities in addition to non-specific arthritis codes to minimize contamination of controls with likely OA subjects (summarized in detail in **Supplementary Table 21**).

### Genotype Data

For both the MVP and the UK Biobank genotype data was generated from DNA typed on a customized Affymetrix Axiom biobank array for both common and rare variants. The MVP samples were genotyped on the Affymetrix Axiom Biobank Array (MVP Release 3). For genotoyped SNPs, standard quality control procedures were implemented to ensure high quality SNP and DNA samples were analyzed^78^. DNA samples were removed for either extreme missingness, gender mismatch, extreme heterozygosity or homozygosity. Imputed results were generated using the 1000 Genomes reference panel and variants with Rsq<0.3 were excluded. A HWE threshold in OA controls in each ancestry and study of p<1E-4 was used to exclude variants. The Harmonizing Genetic Ancestry and Self-Identified Race/Ethnicity (HARE)^66^ approach to classify subjects into major ancestry groups^66^ (see **Supplementary Methods)**. QQ and Manhattan plots for association analyses are provided in Supplementary Materials.

### Trans-ethnic analyses

A challenge with applying traditional meta-analysis methods to diverse populations is that linkage disequilibrium can differ substantially between populations and invalidate the fixed assumption of meta-analyses. For this reason, results from ancestral groups were combined using both a trans-ethnic approach in MANTRA^79^ and a traditional fixed effect in METAL^80^. MANTRA is a meta-analysis method which maximizes homogeneity between closely related populations (i.e. where fixed effect assumption would hold) while allowing for heterogeneity between more diverse groups. The MANTRA approach, described in detail^79^, takes into account the expected similarity in allele effects between more closely related populations without penalizing heterogeneity between more distantly related populations. MANTRA can be conceptualized as a sophisticated method for hybridizing fixed and random effects meta-analyses based on expected allelic similarity of populations. The MANTRA approach has greater power and mapping resolution than random-effects meta-analysis^79^. MANTRA was only used to meta-analyze all 8 populations (**Figure 1C2**: MVP EUR, MVP AFR, MVP ASN, MVP HIS, UKB EUR, UKB AFR, UKB SASN, UKB EASN) and not for study or ancestry stratified meta-analyses (**Figure 1A, 1B**) described above. Fixed effect meta-analyses in METAL were performed for both the overall meta-analysis between all 8 MVP and UKB populations, and the ancestry stratified meta-analyses between MVP and UK Biobank as follows: (1) EUR: MVP EUR, UKB EUR; (2) AFR: MVP AFR, UKB AFR; (3) ASN: MVP ASN, UKB SAS, UKB EAS. For all association results, independent risk SNPs and loci were classified based on LD and block size using FUnctional Mapping and Annotation (FUMA)^81^ software. Specifically, independent SNPs were defined by r2<0.6. Lead independent SNPs were then iteratively defined from the independent SNPs as those with r2<0.1. Whereas independent loci were defined when the distance between independent lead significant SNPs > 250kb.

### Mega GWAS

As illustrated in **Figure 1C1**, we used a linear mixed model approach to perform a cosmopolitan GWAS i.e., not stratifying by or removing subjects who did not cluster with a major ancestry group. We tested the association between each variant and OA using infinitesimal mixed model implemented in BOLT-LMM^17^. Analyses were adjusted for age, sex, BMI and principal components adjusting for population substructure and genetic relatedness in MVP and UKB, separately. Results were combined using fixed effect meta-analysis in METAL. A sensitivity analysis was performed to contrast fixed and random effects meta-analysis with random effect meta-analysis performed using METASOFT^82^.

### Transcriptome-Wide Analyses

Gene-based association tests using MetaXcan^83^ were performed on the meta-analyses created using metal. MetaXcan is machine learning software comprised of tools that integrate genomic information of biological mechanisms with complex traits.

MetaXcan can be used to impute gene-level transcriptomics data in cohorts with only genetic summary statistics using information in a reference set of individuals with both genotype and transcriptomics data. As diverse populations may have different linkage disequilibrium patterns analyses were performed independently. Four tissue-specific prediction models (adipose subcutaneous tissue, brain amygdala, brain frontal cortex, and skeletal muscle) trained on GTEx v8 were chosen for analysis based on their likelihood of providing eQTL information relevant to OA. These tissues were selected based on known factors influencing development of OA such as alterations in anthropometry, physical fitness, and pain sensitivity.

### Fine-mapping and gene-set enrichment analyses

Fine-mapping regions were initially identified using a window (∼50 kb) around the most significant variant; however, we expanded to a wider window where variants’ linkage disequilibrium with the lead variant extended outside the window. This was achieved by manual inspection of regional association plots to ensure the most relevant region was adequately captured. To select five annotation sets for each region, we used the suggested pipeline outlined in the PAINTOR^84-86^ software github repository^87^. Additional details are provided in the **supplementary online methods**. Gene set enrichment analysis (GSEA) ^88^ was performed using FUMA^81^ software.

### Heritability

To calculate heritability we utilized the baseline linkage disequilibrium-linkage disequilibrium adjusted kinships (BLD-LDAK) method which relies on a restricted maximum likelihood (REML) algorithm^89^. SNP summary statistics were used to calculate heritability in respective MVP and UKB.

### Repositionable drugs targeting predicted OA induced gene expression

We tested whether significant genes identified via transcriptome-wide imputation are enriched with targets for drug repurposing using Genome for REPositioning drugs (GREP)^90^ software. The set of genes predicted to be eQTL (P<0.05) in MetaXcan of the mega-GWAS results was input to GREP. The method performs Fisher’s exact tests to test whether input genes are enriched with genes targeted by drugs with clinical indication category in the Anatomical Therapeutic Chemical Classification System (ATC).

## Funding

This research is based on data from the Million Veteran Program, Office of Research and Development, Veterans Health Administration, and was supported by award #I01RX002745. This publication does not represent the views of the Department of Veteran Affairs or the United States Government.

## Conflicts of Interest

Jasvinder A. Singh has received consultant fees from Crealta/Horizon, Medisys, Fidia, UBM LLC, Trio health, Medscape, WebMD, PK Med, Two labs Inc., Adept Field Solutions, Clinical Care options, Clearview healthcare partners, Putnam associates, Focus forward, Navigant consulting, Spherix, Practice Point communications, the National Institutes of Health and the American College of Rheumatology. Jasvinder A. Singh has received institutional research support from Zimmer Biomet Holdings. Jasvinder A. Singh received food and beverage payments from Intuitive Surgical Inc./Philips Electronics North America. Jasvinder A. Singh owns stock options in TPT Global Tech, Vaxart pharmaceuticals, Atyu biopharma, Adaptimmune Therapeutics, GeoVax Labs, Pieris Pharmaceuticals, Enzolytics Inc., Seres Therapeutics, Tonix Pharmaceuticals Holding Corp., and Charlotte’s Web Holdings, Inc. Jasvinder A. Singh previously owned stock options in Amarin, Viking and Moderna pharmaceuticals. Jasvinder A. Singh is on the speaker’s bureau of Simply Speaking. Jasvinder A. Singh is a member of the executive of Outcomes Measures in Rheumatology (OMERACT), an organization that develops outcome measures in rheumatology and receives arms-length funding from 12 companies. Jasvinder A. Singh serves on the FDA Arthritis Advisory Committee. Jasvinder A. Singh is the chair of the Veterans Affairs Rheumatology Field Advisory Committee. Jasvinder A. Singh is the editor and the Director of the University of Alabama at Birmingham (UAB) Cochrane Musculoskeletal Group Satellite Center on Network Meta-analysis. Jasvinder A. Singh previously served as a member of the following committees: member, the American College of Rheumatology’s (ACR) Annual Meeting Planning Committee (AMPC) and Quality of Care Committees, the Chair of the ACR Meet-the-Professor, Workshop and Study Group Subcommittee and the co-chair of the ACR Criteria and Response Criteria subcommittee.

## McDonald Lab at the University of Alabama at Birmingham (UAB)

Rachel Tindal, medical student, for assistance with manuscript editing. Cameron Cole, summer student, for assistance formatting data.

## VA Million Veteran Program: Core Acknowledgement

### MVP Executive Committee

- Co-Chair: J. Michael Gaziano, M.D., M.P.H. VA Boston Healthcare System, 150 S. Huntington Avenue, Boston, MA 02130
- Co-Chair: Sumitra Muralidhar, Ph.D. US Department of Veterans Affairs, 810 Vermont Avenue NW, Washington, DC 20420
- Rachel Ramoni, D.M.D., Sc.D., Chief VA Research and Development Officer US Department of Veterans Affairs, 810 Vermont Avenue NW, Washington, DC 20420
- Jean Beckham, Ph.D. Durham VA Medical Center, 508 Fulton Street, Durham, NC 27705
- Kyong-Mi Chang, M.D. Philadelphia VA Medical Center, 3900 Woodland Avenue, Philadelphia, PA 19104
- Christopher J. O’Donnell, M.D., M.P.H. VA Boston Healthcare System, 150 S. Huntington Avenue, Boston, MA 02130
- Philip S. Tsao, Ph.D. VA Palo Alto Health Care System, 3801 Miranda Avenue, Palo Alto, CA 94304
- James Breeling, M.D., Ex-Officio US Department of Veterans Affairs, 810 Vermont Avenue NW, Washington, DC 20420
- Grant Huang, Ph.D., Ex-Officio US Department of Veterans Affairs, 810 Vermont Avenue NW, Washington, DC 20420
- Juan P. Casas, M.D., Ph.D., Ex-Officio VA Boston Healthcare System, 150 S. Huntington Avenue, Boston, MA 02130

### MVP Program Office

- Sumitra Muralidhar, Ph.D. US Department of Veterans Affairs, 810 Vermont Avenue NW, Washington, DC 20420
- Jennifer Moser, Ph.D. US Department of Veterans Affairs, 810 Vermont Avenue NW, Washington, DC 20420

### MVP Recruitment/Enrollment

- Recruitment/Enrollment Director/Deputy Director, Boston – Stacey B. Whitbourne, Ph.D.; Jessica V. Brewer, M.P.H. VA Boston Healthcare System, 150 S. Huntington Avenue, Boston, MA 02130
- MVP Coordinating Centers
  ○ Clinical Epidemiology Research Center (CERC), West Haven – Mihaela Aslan, Ph.D. West Haven VA Medical Center, 950 Campbell Avenue, West Haven, CT 06516
  ○ Cooperative Studies Program Clinical Research Pharmacy Coordinating Center, Albuquerque – Todd Connor, Pharm.D.; Dean P. Argyres, B.S., M.S. New Mexico VA Health Care System, 1501 San Pedro Drive SE, Albuquerque, NM 87108
  ○ Genomics Coordinating Center, Palo Alto – Philip S. Tsao, Ph.D. VA Palo Alto Health Care System, 3801 Miranda Avenue, Palo Alto, CA 94304
  ○ MVP Boston Coordinating Center, Boston-J. Michael Gaziano, M.D., M.P.H. VA Boston Healthcare System, 150 S. Huntington Avenue, Boston, MA 02130
  ○ MVP Information Center, Canandaigua – Brady Stephens, M.S. Canandaigua VA Medical Center, 400 Fort Hill Avenue, Canandaigua, NY 14424
- VA Central Biorepository, Boston – Mary T. Brophy M.D., M.P.H.; Donald E. Humphries, Ph.D.; Luis E. Selva, Ph.D. VA Boston Healthcare System, 150 S. Huntington Avenue, Boston, MA 02130
- MVP Informatics, Boston – Nhan Do, M.D.; Shahpoor (Alex) Shayan, M.S. VA Boston Healthcare System, 150 S. Huntington Avenue, Boston, MA 02130
- MVP Data Operations/Analytics, Boston – Kelly Cho, M.P.H., Ph.D. VA Boston Healthcare System, 150 S. Huntington Avenue, Boston, MA 02130
- Director of Regulatory Affairs – Lori Churby, B.S. VA Palo Alto Health Care System, 3801 Miranda Avenue, Palo Alto, CA 94304

### MVP Science

- Science Operations – Christopher J. O’Donnell, M.D., M.P.H. VA Boston Healthcare System, 150 S. Huntington Avenue, Boston, MA 02130
- Genomics Core – Christopher J. O’Donnell, M.D., M.P.H.; Saiju Pyarajan Ph.D. VA Boston Healthcare System, 150 S. Huntington Avenue, Boston, MA 02130 Philip S. Tsao, Ph.D. VA Palo Alto Health Care System, 3801 Miranda Avenue, Palo Alto, CA 94304
- Data Core – Kelly Cho, M.P.H, Ph.D. VA Boston Healthcare System, 150 S. Huntington Avenue, Boston, MA 02130
- VA Informatics and Computing Infrastructure (VINCI) – Scott L. DuVall, Ph.D. VA Salt Lake City Health Care System, 500 Foothill Drive, Salt Lake City, UT 84148
- Data and Computational Sciences – Saiju Pyarajan, Ph.D. VA Boston Healthcare System, 150 S. Huntington Avenue, Boston, MA 02130
- Statistical Genetics – Elizabeth Hauser, Ph.D. Durham VA Medical Center, 508 Fulton Street, Durham, NC 27705 Yan Sun, Ph.D.
- Atlanta VA Medical Center, 1670 Clairmont Road, Decatur, GA 30033 Hongyu Zhao, Ph.D. West Haven VA Medical Center, 950 Campbell Avenue, West Haven, CT 06516

### Current MVP Local Site Investigators

- Atlanta VA Medical Center (Peter Wilson, M.D.) 1670 Clairmont Road, Decatur, GA 30033
- Bay Pines VA Healthcare System (Rachel McArdle, Ph.D.) 10,000 Bay Pines Blvd Bay Pines, FL 33744
- Birmingham VA Medical Center (Louis Dellitalia, M.D.) 700 S. 19th Street, Birmingham AL 35233
- Central Western Massachusetts Healthcare System (Kristin Mattocks, Ph.D., M.P.H.) 421 North Main Street, Leeds, MA 01053
- Cincinnati VA Medical Center (John Harley, M.D., Ph.D.) 3200 Vine Street, Cincinnati, OH 45220
- Clement J. Zablocki VA Medical Center (Jeffrey Whittle, M.D., M.P.H.) 5000 West National Avenue, Milwaukee, WI 53295
- VA Northeast Ohio Healthcare System (Frank Jacono, M.D.) 10701 East Boulevard, Cleveland, OH 44106
- Durham VA Medical Center (Jean Beckham, Ph.D.) 508 Fulton Street, Durham, NC 27705
- Edith Nourse Rogers Memorial Veterans Hospital (John Wells., Ph.D.) 200 Springs Road, Bedford, MA 01730
- Edward Hines, Jr. VA Medical Center (Salvador Gutierrez, M.D.) 5000 South 5th Avenue, Hines, IL 60141
- Veterans Health Care System of the Ozarks (Gretchen Gibson, D.D.S., M.P.H.) 1100 North College Avenue, Fayetteville, AR 72703
- Fargo VA Health Care System (Kimberly Hammer, Ph.D.) 2101 N. Elm, Fargo, ND 58102
- VA Health Care Upstate New York (Laurence Kaminsky, Ph.D.) 113 Holland Avenue, Albany, NY 12208
- New Mexico VA Health Care System (Gerardo Villareal, M.D.) 1501 San Pedro Drive, S.E. Albuquerque, NM 87108
- VA Boston Healthcare System (Scott Kinlay, M.B.B.S., Ph.D.) 150 S. Huntington Avenue, Boston, MA 02130
- VA Western New York Healthcare System (Junzhe Xu, M.D.) 3495 Bailey Avenue, Buffalo, NY 14215-1199
- Ralph H. Johnson VA Medical Center (Mark Hamner, M.D.) 109 Bee Street, Mental Health Research, Charleston, SC 29401
- Columbia VA Health Care System (Roy Mathew, M.D.) 6439 Garners Ferry Road, Columbia, SC 29209
- VA North Texas Health Care System (Sujata Bhushan, M.D.) 4500 S. Lancaster Road, Dallas, TX 75216
- Hampton VA Medical Center (Pran Iruvanti, D.O., Ph.D.) 100 Emancipation Drive, Hampton, VA 23667
- Richmond VA Medical Center (Michael Godschalk, M.D.) 1201 Broad Rock Blvd., Richmond, VA 23249
- Iowa City VA Health Care System (Zuhair Ballas, M.D.) 601 Highway 6 West, Iowa City, IA 52246-2208
- Eastern Oklahoma VA Health Care System (Douglas Ivins, M.D.) 1011 Honor Heights Drive, Muskogee, OK 74401
- James A. Haley Veterans’ Hospital (Stephen Mastorides, M.D.) 13000 Bruce B. Downs Blvd, Tampa, FL 33612
- James H. Quillen VA Medical Center (Jonathan Moorman, M.D., Ph.D.) Corner of Lamont & Veterans Way, Mountain Home, TN 37684
- John D. Dingell VA Medical Center (Saib Gappy, M.D.) 4646 John R Street, Detroit, MI 48201
- Louisville VA Medical Center (Jon Klein, M.D., Ph.D.) 800 Zorn Avenue, Louisville, KY 40206
- Manchester VA Medical Center (Nora Ratcliffe, M.D.) 718 Smyth Road, Manchester, NH 03104
- Miami VA Health Care System (Hermes Florez, M.D., Ph.D.) 1201 NW 16th Street, 11 GRC, Miami FL 33125
- Michael E. DeBakey VA Medical Center (Olaoluwa Okusaga, M.D.) 2002 Holcombe Blvd, Houston, TX 77030
- Minneapolis VA Health Care System (Maureen Murdoch, M.D., M.P.H.) One Veterans Drive, Minneapolis, MN 55417
- N. FL/S. GA Veterans Health System (Peruvemba Sriram, M.D.) 1601 SW Archer Road, Gainesville, FL 32608
- Northport VA Medical Center (Shing Shing Yeh, Ph.D., M.D.) 79 Middleville Road, Northport, NY 11768
- Overton Brooks VA Medical Center (Neeraj Tandon, M.D.) 510 East Stoner Ave, Shreveport, LA 71101
- Philadelphia VA Medical Center (Darshana Jhala, M.D.) 3900 Woodland Avenue, Philadelphia, PA 19104
- Phoenix VA Health Care System (Samuel Aguayo, M.D.) 650 E. Indian School Road, Phoenix, AZ 85012
- Portland VA Medical Center (David Cohen, M.D.) 3710 SW U.S. Veterans Hospital Road, Portland, OR 97239
- Providence VA Medical Center (Satish Sharma, M.D.) 830 Chalkstone Avenue, Providence, RI 02908
- Richard Roudebush VA Medical Center (Suthat Liangpunsakul, M.D., M.P.H.) 1481 West 10th Street, Indianapolis, IN 46202
- Salem VA Medical Center (Kris Ann Oursler, M.D.) 1970 Roanoke Blvd, Salem, VA 24153
- San Francisco VA Health Care System (Mary Whooley, M.D.) 4150 Clement Street, San Francisco, CA 94121
- South Texas Veterans Health Care System (Sunil Ahuja, M.D.) 7400 Merton Minter Boulevard, San Antonio, TX 78229
- Southeast Louisiana Veterans Health Care System (Joseph Constans, Ph.D.) 2400 Canal Street, New Orleans, LA 70119
- Southern Arizona VA Health Care System (Paul Meyer, M.D., Ph.D.) 3601 S 6th Avenue, Tucson, AZ 85723
- Sioux Falls VA Health Care System (Jennifer Greco, M.D.) 2501 W 22nd Street, Sioux Falls, SD 57105
- St. Louis VA Health Care System (Michael Rauchman, M.D.) 915 North Grand Blvd, St. Louis, MO 63106
- Syracuse VA Medical Center (Richard Servatius, Ph.D.) 800 Irving Avenue, Syracuse, NY 13210
- VA Eastern Kansas Health Care System (Melinda Gaddy, Ph.D.) 4101 S 4th Street Trafficway, Leavenworth, KS 66048
- VA Greater Los Angeles Health Care System (Agnes Wallbom, M.D., M.S.) 11301 Wilshire Blvd, Los Angeles, CA 90073
- VA Long Beach Healthcare System (Timothy Morgan, M.D.) 5901 East 7th Street Long Beach, CA 90822
- VA Maine Healthcare System (Todd Stapley, D.O.) 1 VA Center, Augusta, ME 04330
- VA New York Harbor Healthcare System (Scott Sherman, M.D., M.P.H.) 423 East 23rd Street, New York, NY 10010
- VA Pacific Islands Health Care System (George Ross, M.D.) 459 Patterson Rd, Honolulu, HI 96819
- VA Palo Alto Health Care System (Philip Tsao, Ph.D.) 3801 Miranda Avenue, Palo Alto, CA 94304-1290
- VA Pittsburgh Health Care System (Patrick Strollo, Jr., M.D.) University Drive, Pittsburgh, PA 15240
- VA Puget Sound Health Care System (Edward Boyko, M.D.) 1660 S. Columbian Way, Seattle, WA 98108-1597
- VA Salt Lake City Health Care System (Laurence Meyer, M.D., Ph.D.) 500 Foothill Drive, Salt Lake City, UT 84148
- VA San Diego Healthcare System (Samir Gupta, M.D., M.S.C.S.) 3350 La Jolla Village Drive, San Diego, CA 92161
- VA Sierra Nevada Health Care System (Mostaqul Huq, Pharm.D., Ph.D.) 975 Kirman Avenue, Reno, NV 89502
- VA Southern Nevada Healthcare System (Joseph Fayad, M.D.) 6900 North Pecos Road, North Las Vegas, NV 89086
- VA Tennessee Valley Healthcare System (Adriana Hung, M.D., M.P.H.) 1310 24th Avenue, South Nashville, TN 37212
- Washington DC VA Medical Center (Jack Lichy, M.D., Ph.D.) 50 Irving St, Washington, D. C. 20422
- W.G. (Bill) Hefner VA Medical Center (Robin Hurley, M.D.) 1601 Brenner Ave, Salisbury, NC 28144
- White River Junction VA Medical Center (Brooks Robey, M.D.) 163 Veterans Drive, White River Junction, VT 05009
- William S. Middleton Memorial Veterans Hospital (Robert Striker, M.D., Ph.D.) 2500 Overlook Terrace, Madison, WI 53705

