## Supplementary Materials for "Novel Genetic Loci Associated with Osteoarthritis in Multi-Ancestry Analyses in 484,374 Participants from MVP and the UK Biobank"

**SUPPLEMENTARY MATERIAL**

**ONLINE METHODS**

Approval for the study and the work described in this manuscript received ethical and study protocol approval from the Veteran Affair Central Institutional Review Board as well as the University of Alabama at Birmingham in accordance with the principles outlined in the Declaration of Helsinki.

**Million Veteran Program**: The study has been previously described in detail[1]. The current analyses included participants between 40 and 80 years of age, although the MVP recruited subjects between 19-104 years from 50 Veterans Affairs Medical Centers nationwide since 2011. Each Veteran participant's electronic health record (EHR) data as well as additional completed MVP questionnaire data was integrated into the MVP biorepository

**UK Biobank**: The UK Biobank (UKB) is an open-access resource with phenotype and genotype data from 500,000 volunteer participants recruited from the United Kingdom that is available to approved researchers from academia and industry[39]. The UKB is prospective cohort that has extensive phenotypic information and has generated genotype data using the UKB Axiom Array, very similar to the MVP Axiom Array. Relevant longitudinal EHR data and additional questionnaire data administered at baseline enrollment are available. UK Biobank data were analyzed under approved access via proposal #30350.

**Osteoarthritis trait definition:** Osteoarthritis cases and controls were identified using International Classification of Disease, 9$^{th}$ revision, common modification (ICD-9-CM) and ICD-10-CM codes from the MVP diagnosis file version 18.2. Diagnosis dates ranged from July 2, 1990 to June 13, 2019. ICD code lists from Zengini et al. [2] were used. For OA case identification, lists of codes were provided for a general OA category and overlapping lists for specific subtypes of OA (Hip, Knee, Spine, Hand, Finger, and Thumb) provided in **Supplementary Table 20**. Separate lists were also provided for exclusion criteria for controls which included a wide range of non-OA arthropathies detailed in **Supplementary Table 21.** We further identified a list of codes for traumatic and post-traumatic arthropathy by examining ICD9 and ICD10 code descriptions for the string 'trauma' and identifying those denoting traumatic or post-traumatic arthropathy. These included ICD9-CM codes '716.1, 716.10, 716.11, 716.12, 716.13, 716.14, 716.15, 716.16, 716.17, 716.18, 716.19' and ICD10-CM codes 'M12.50, M12.519, M12.529, M12.539, M12.549, M12.559, M12.569, M12.579, M12.58, M12.59, M19.1, M19.10, M19.11, M19.12, M19.13, M19.14, M19.15, M19.16, M19.17, M19.18, M19.19'. Of these, the list 716.10, M12.50,

M12.519, M12.529, M12.539, M12.549, M12.559, M12.569, M12.579, M12.58, M12.59 were not included in the list of control-exclusions.

To increase specificity, OA cases were required to have at least two codes within a category at least 30 days apart regardless of category (e.g. one general OA code and one Spine OA code at least 30 days apart). Cases of specific OA subtypes were identified by being an OA case and having a single code within the subtype, thus a person with a single code for Hip OA and another code for Hand OA would be counted as having OA (**Supplementary Figure 1**).  A similar process was undertaken to identify OA cases and controls in the UK Biobank but without requiring that OA cases have two or more codes 30 days apart because date of ICD code was not provided; only occurrence of the code (**Supplementary Figure 1**).

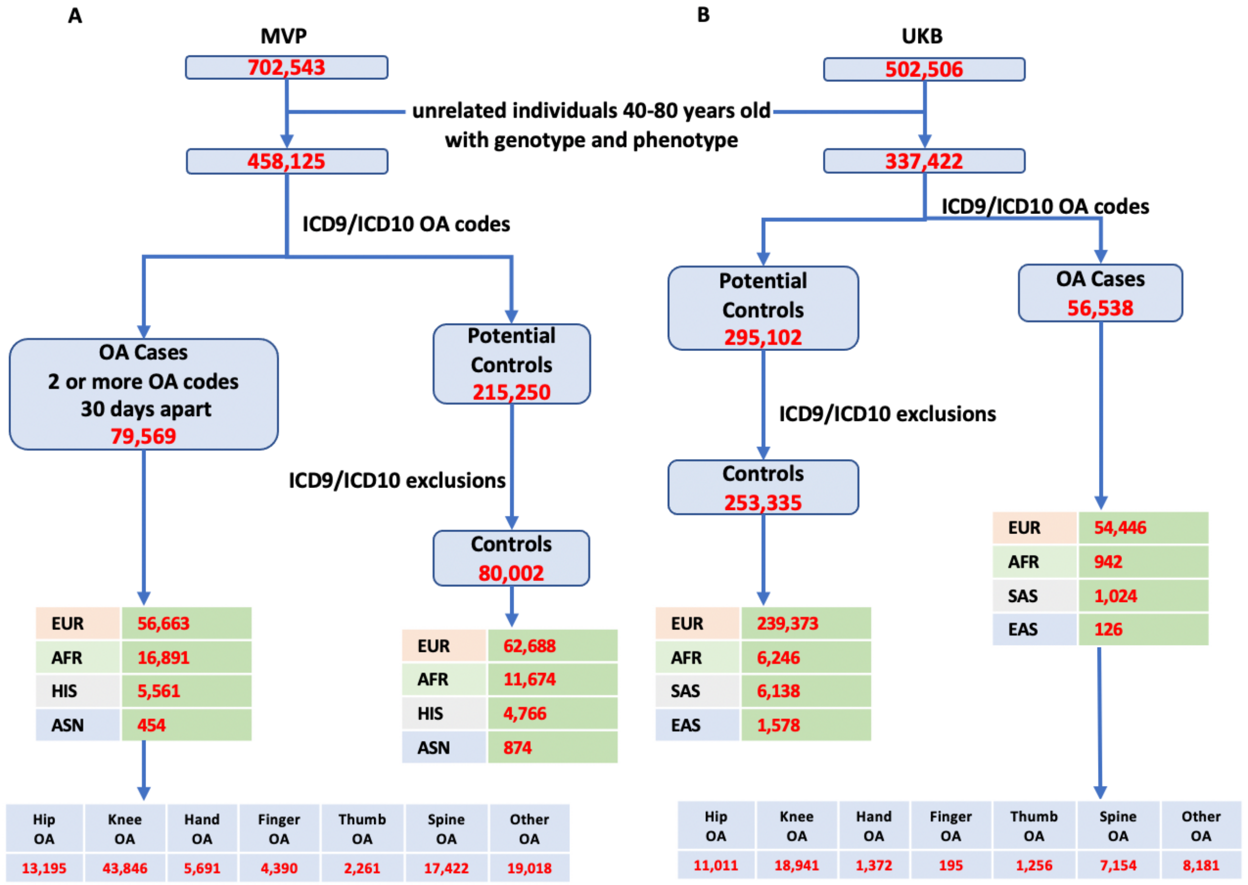


**Supplementary Figure 1:** Coding of OA cases and controls in the Million Veteran Project  (A) and the UK Biobank (B). EUR: European White, AFR: African, HIS: Hispanic, ASN: Asian, SAS: South Asian, EAS: East Asian descent.

**Classification of UKB subjects into major ancestry groups:** We used the Harmonizing Genetic Ancestry and Self-Identified Race/Ethnicity (HARE)[3] approach to classify UKB subjects. This approach is an expansion of methods that use genetic data to characterize race by also harmonizing self-reported race and ethnicity in the classification algorithm. Unrelated UKB subjects with self-identified race/ethnicity (SIRE) information were categorized into 4 major ancestral groups: European White (EUR), African (AFR), East Asian (EAS), or South Asian (SAS). A support vector machine (SVM) model was built based on correspondence between Genetically Inferred Ancestry (GIA) defined by the top 30 principal components (PCs) accounting for population structure and SIRE. The SVM was run using default tuning parameters, t1=20 and t2=40, which control the stringencies with which the outliers are removed and individuals without SIRE are assigned a HARE. This led to HARE classification of 294,694 EUR, 7,300 AFR, 7,240 SAS and 1,715 EAS subjects in the UKB (**Supplementary Figure 2**).

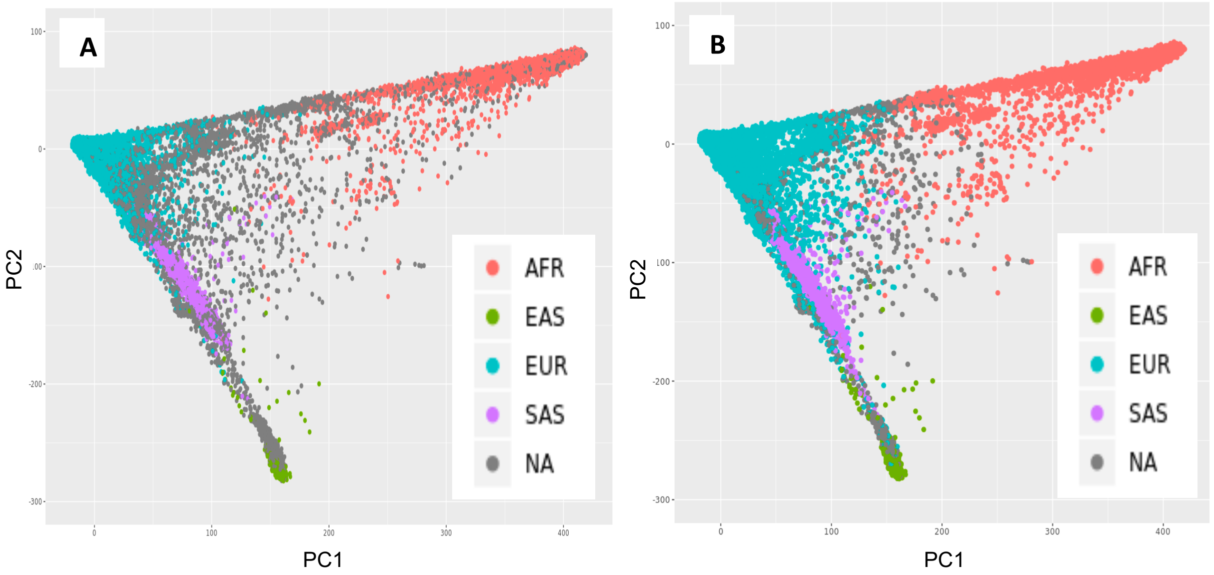


**Supplementary Figure 2: First two Principal Components (PCs) of Genetically Inferred Ancestry (GIA) with Self Identified Race Ethnicity (SIRE) and Harmonizing Ancestry and Race/Ethnicity (HARE) assignments in the UKB.** Panel **A** displays relationship between GIA PCs with initial SIRE. Panel **B** displays relationship between GIA PCs with final HARE assignments.

**Association testing within ancestral groups**: Genetic variants were tested for association with OA assuming an additive model adjusting for age, sex and BMI in addition to the top 10 PCs in each ancestral group and was used to adjust for population substructure using PLINK software. Results were meta-analyzed using fixed effect meta-analysis in METAL as well as using MANTRA[4], a meta-analysis method developed for trans-ethnic fine mapping, which maximizes homogeneity between closely related populations while allowing for heterogeneity between more diverse groups.

**Transcriptome-Wide Analyses:** Gene-based association tests using MetaXcan were performed on the meta-analyses results generated using metal. MetaXcan is a machine learning software comprised of tools that integrate genomic information of biological mechanisms with complex traits. MetaXcan can be used to impute gene-level transcriptomics data in cohorts with only genetic summary statistics using information in a reference set of individuals with both genotype and transcriptomics data. As populations of African, European, and Asian descent may have different linkage disequilibrium patterns analyses were performed independently. Four tissue specific prediction models (adipose subcutaneous tissue, brain amygdala, brain frontal cortex and skeletal muscle trained on GTEx v8 were chosen for analysis based on their likelihood of providing eQTL information relevant to OA.

**Fine-mapping:** Fine-mapping regions were initially identified using a window around the most significant variant. Manual inspection of regional association plots was also performed to ensure the most relevant region was adequately captured. We used the iterative procedure outlined in the PAINTOR manuscript as well as in **Supplementary Figure 3** to select annotation sets for each region.

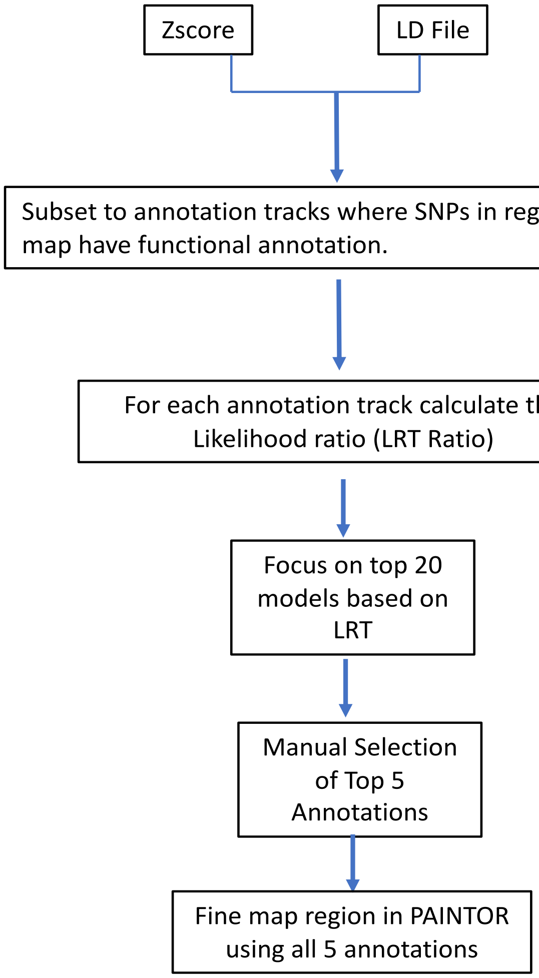


**Supplementary Figure 3**. PAINTOR fine-mapping pipeline.

**Heritability:** In this study, SNP heritability ($h^2_{SNP}$), defined as the proportion of total phenotypic variance due to additive genetic effects of a given set of SNPs, was used to estimate the heritability of the binary OA phenotype from GWAS summary statistics in the MVP and UKBiobank cohorts. To calculate heritability, SNPs were filtered from each GWAS analysis on the basis of an imputation r-squared value > 0.8, 0.005 < MAF < 0.95, excluding indels, structural variants, and strand-ambiguous SNPs. To calculate heritability we utilized the baseline linkage disequilibrium-linkage disequilibrium adjusted kinships (BLD-LDAK) method which relies on a restricted maximum likelihood (REML) algorithm[1]. In brief, the BLD-LDAK model accounts for the potential bias of assuming an equal, additive effect of each SNP to heritability estimates through a re-weighting of SNPs to account for LD and quality. Thus, higher quality SNPs in low LD regions contribute more than lower quality SNPs in high LD regions. In addition, the BLD-LDAK model allows for the use of a tagging file containing annotated groups of SNPs for functional groups of interest (i.e., coding regions, enhancers, among others) that can be used to estimate what percent each functional group contributes to the overall measure of heritability. In our study, tagging files were created for each ancestry using genotype data from the 1000 Genomes Project as a reference panel[2]. In addition to the 66 annotated groups provided by BLD-LDAK, two additional annotation groups were created for each ancestry in MVP and UKB to denote OA eQTLs, which were identified as SNPs with a p<0.001 from MANTRA analyses. In case-control studies, heritability can be biased by disease prevalence in the population. To account for this potential bias, a population prevalence of 10% and case frequencies were incorporated into the BLD-LDAK model to obtain robust heritability estimations.

**SUPPLEMENTARY RESULTS**

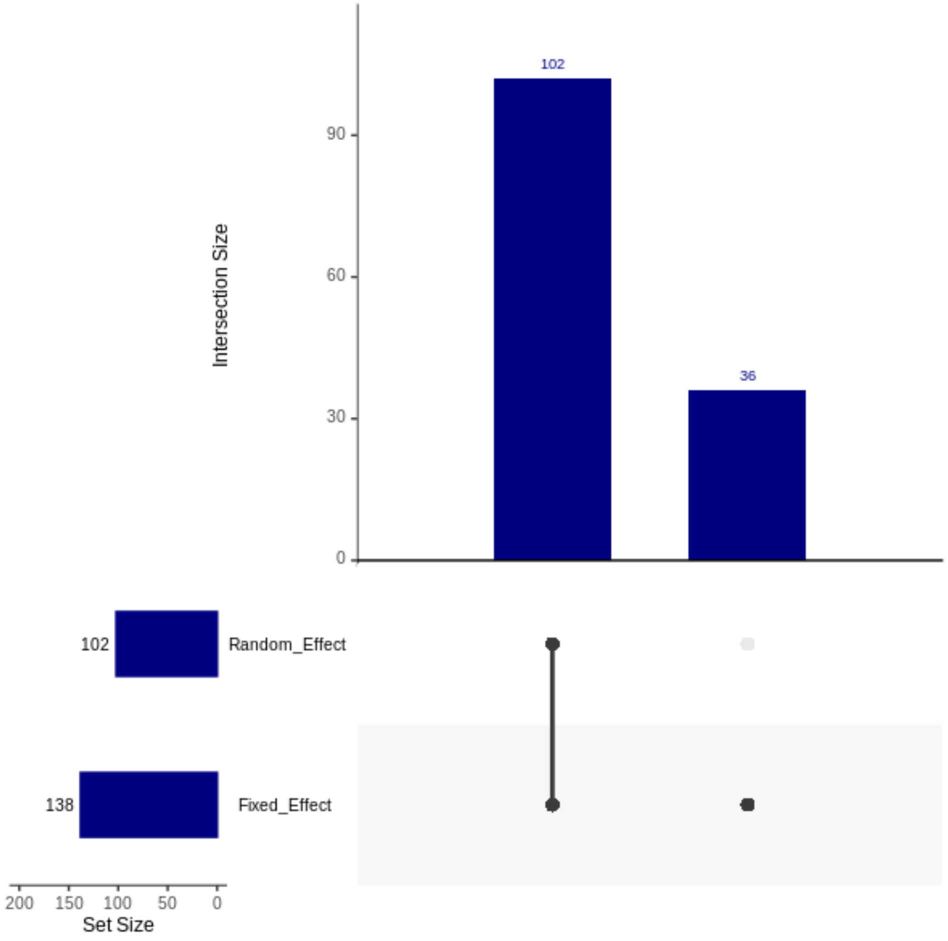


**Supplementary Figure 4:** Sensitivity analysis comparing concordance of significant SNPs associated with OA in MVP using fixed and/or random-effects meta-analyses methods. Analysis indicates no additional variants are captured using random effects meta-analysis.

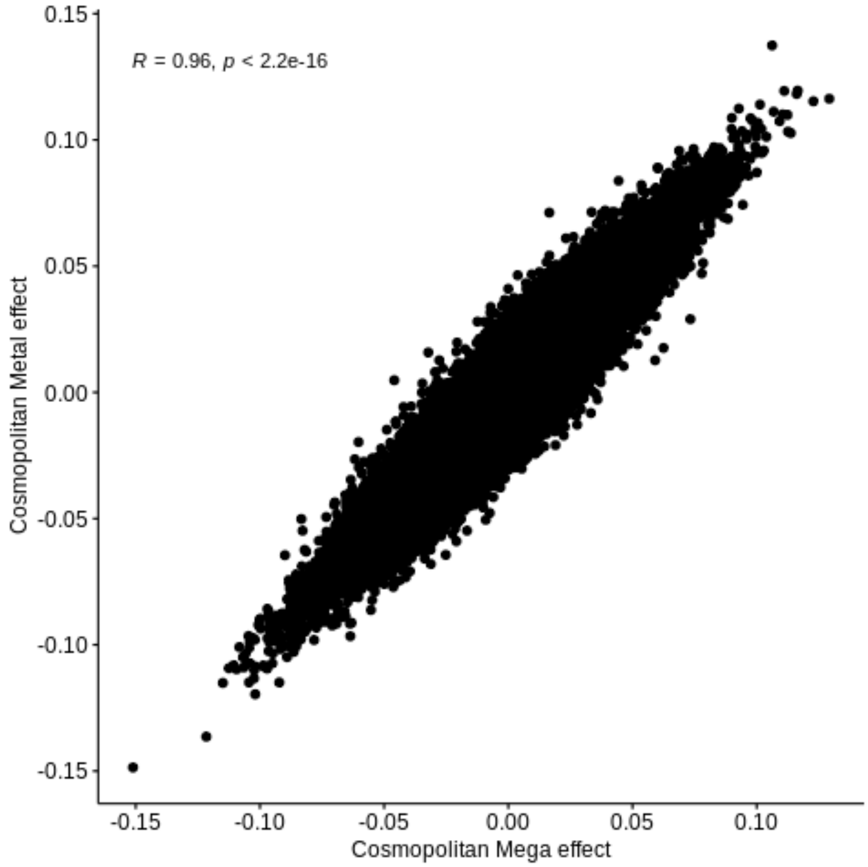


**Supplementary Figure 5:** Correlation plot of the SNP association effect-size for the Mega and fixed effect meta-analyses approaches.

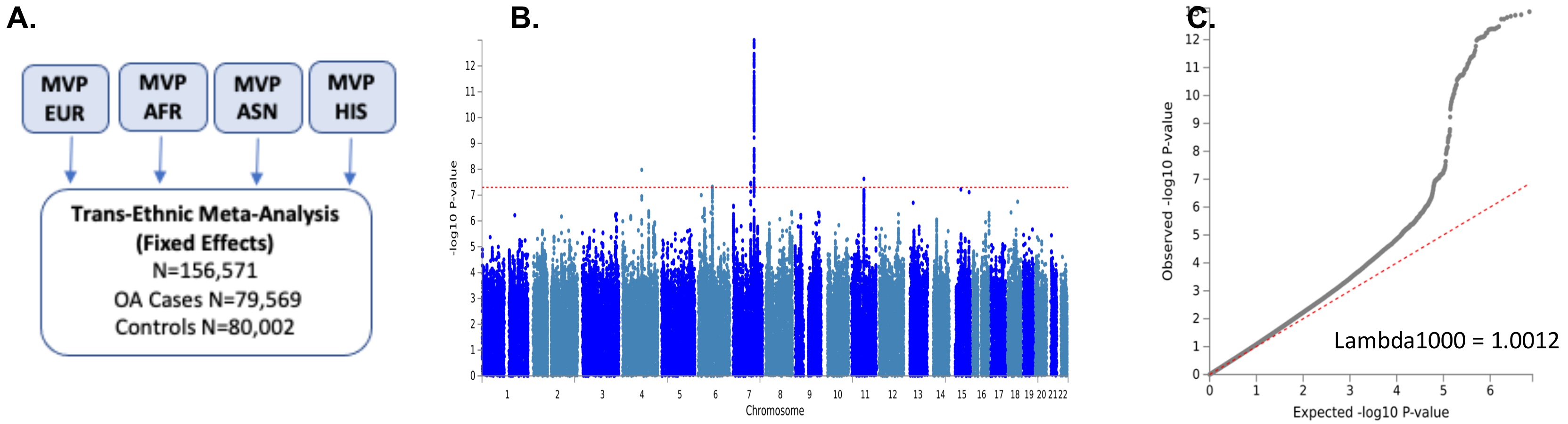


**Supplementary Figure 6:** Manhattan and QQ plots of trans-ethnic meta-analysis variants associated with OA in MVP. A. GWAS result study design. B. Manhattan plot. C. QQ plot.

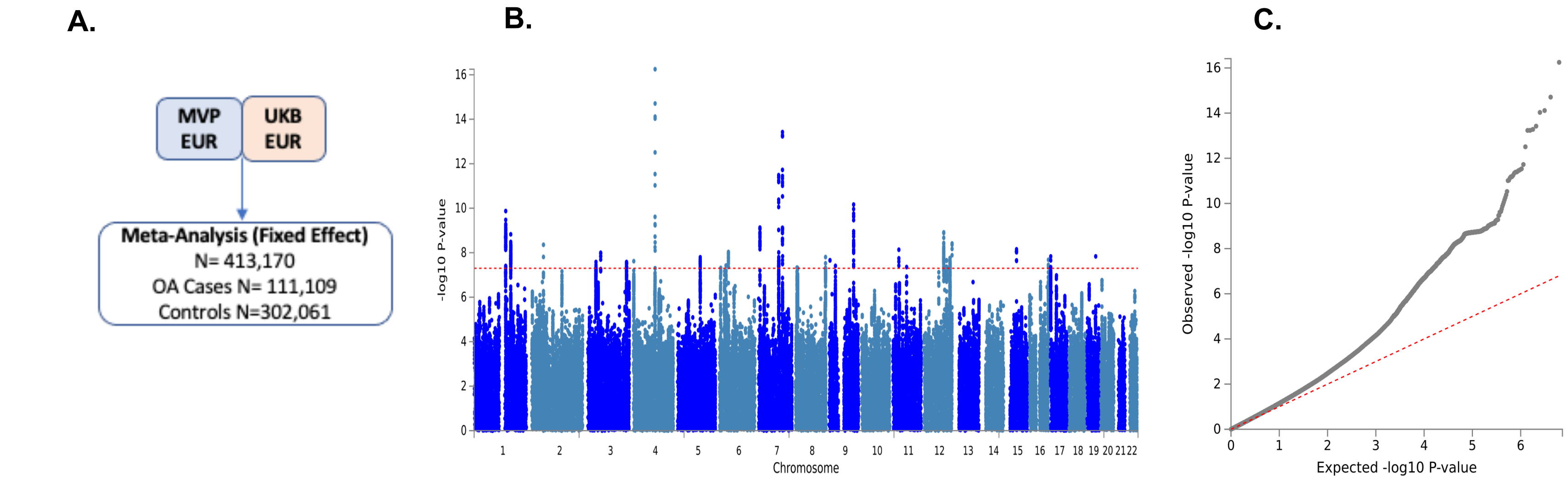


**Supplementary Figure 7:** Manhattan and QQ plots of fixed effect meta-analysis of variants associated with OA in participants of EUR descent in MVP and UKB. A. GWAS result study design. B. Manhattan plot. C. QQ plot.

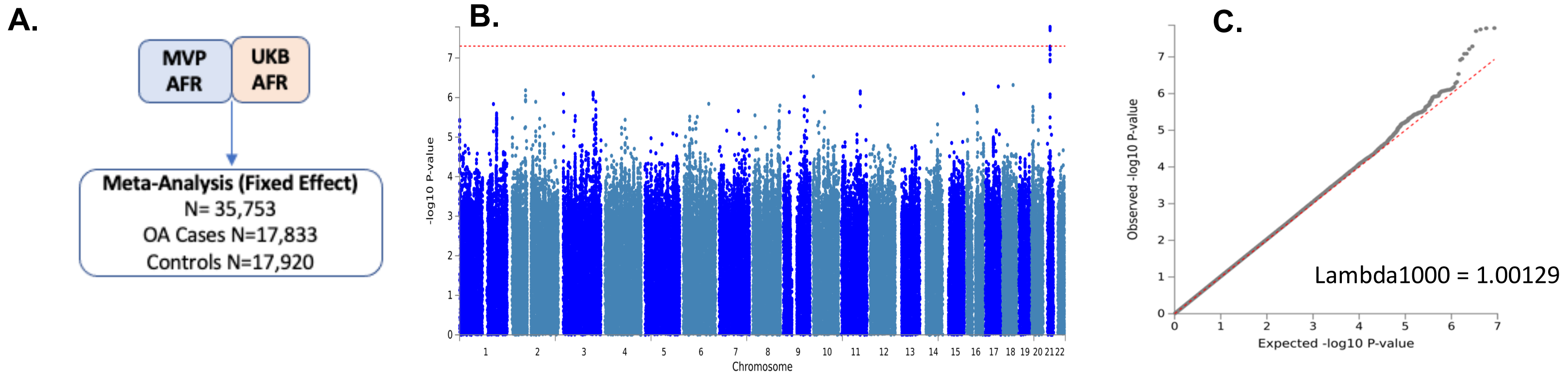


**Supplementary Figure 8:** Manhattan and QQ plots of fixed effect meta-analysis of variants associated with OA in participants of AFR descent in MVP and UKB. A. GWAS result study design. B. Manhattan plot. C. QQ plot.

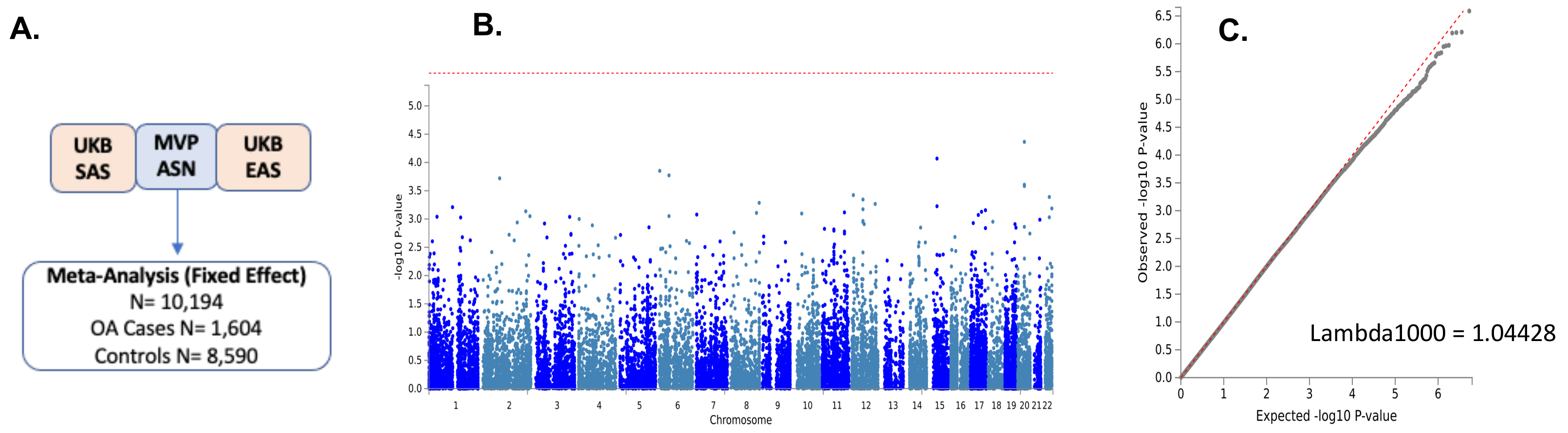


**Supplementary Figure 9:** Manhattan and QQ plots of fixed effect meta-analysis of variants associated with OA in participants of ASN descent in MVP and UKB. A. GWAS result study design. B. Manhattan plot. C. QQ plot.

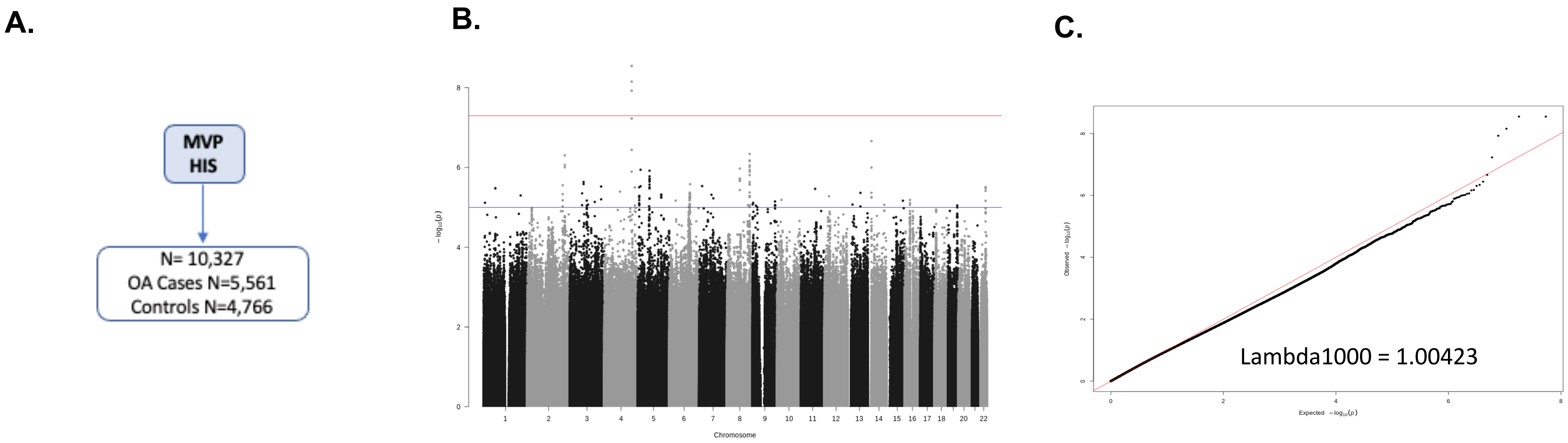


**Supplementary Figure 10:** Manhattan and QQ plots of variants associated with OA in Veterans of HIS descent in MVP. A. GWAS result study design. B. Manhattan plot. C. QQ plot.

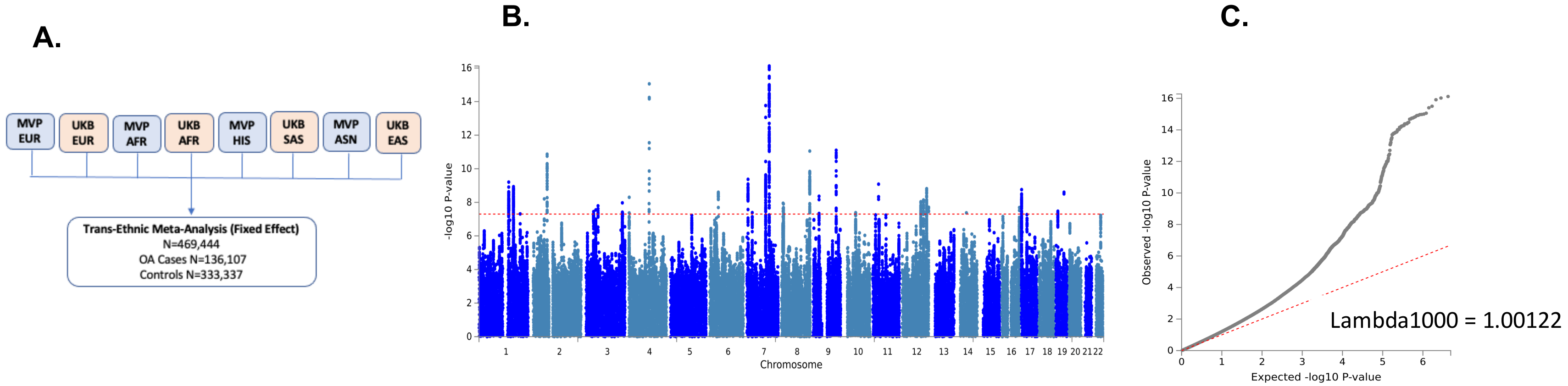


**Supplementary Figure 11:** Manhattan and QQ plots of trans-ethnic fixed effect meta-analysis of variants associated with OA in participants from MVP and UKB. A. GWAS result study design. B. Manhattan plot. C. QQ plot.

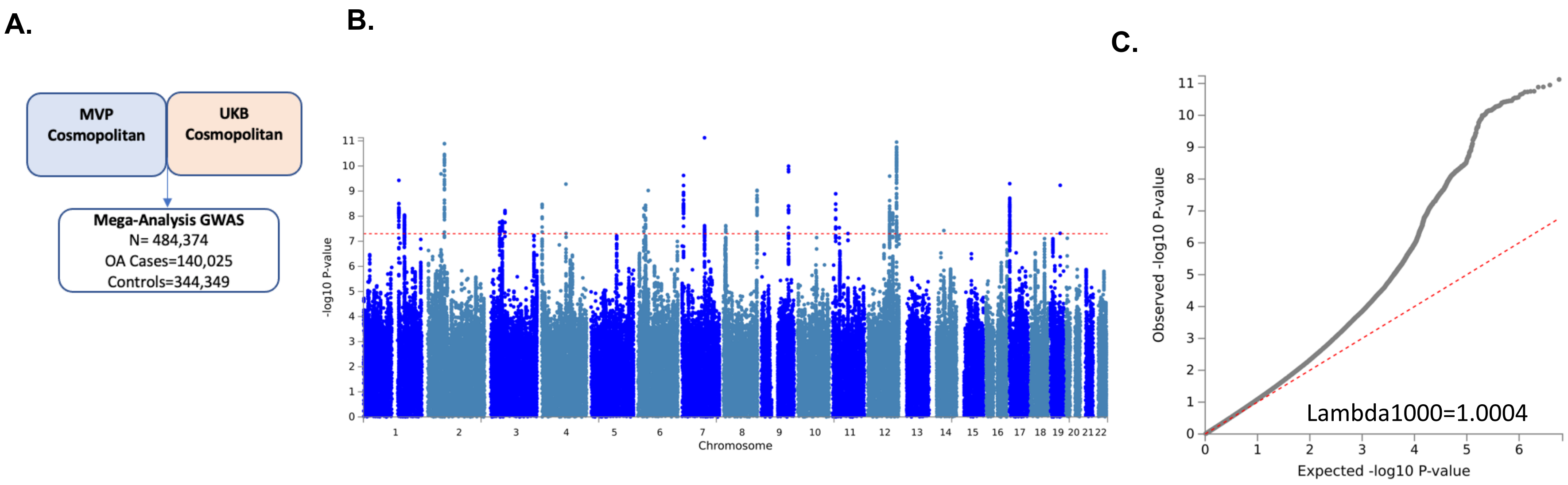


**Supplementary Figure 12:** Manhattan and QQ plots of mega-analysis of variants associated with OA in participants from MVP and UKB. A. GWAS result study design. B. Manhattan plot. C. QQ plot.
